# Tracking Cytopenias in *FANCA*-deficient Fanconi Anemia

**DOI:** 10.64898/2025.12.14.25341731

**Authors:** Rochelle R. Maxwell, Tamar Berger, Caroline S. Jiang, Allana Rosenberg, Ashlyn-Maree Gonzalez, Jodie Odame, Yu-Chien Lin, Francis P. Lach, Jennifer Kennedy, Rebecca Tryon, Frank X. Donovan, Danielle C. Kimble, Shivatheja Soma, Maria I. Cancio, John E. Wagner, Margaret L. MacMillan, Stella M. Davies, Settara C. Chandrasekharappa, Parinda A. Mehta, Farid Boulad, Arleen D. Auerbach, Agata Smogorzewska

## Abstract

Fanconi anemia (FA) is an inherited disorder classically characterized by childhood-onset bone marrow dysfunction and lifelong cancer predisposition. FA is caused by pathogenic variants in any one of 23 genes identified so far. Of these, *FANCA* is the most frequently mutated and accounts for disease in two-thirds of all patients with FA. The spectrum of *FANCA* pathogenic variants (mutations) is broad, and genotype-phenotype correlation is often unclear. Here we describe the natural history of cytopenias associated with *FANCA* pathogenic variants in 139 individuals diagnosed in 1995 or later. We followed blood cell counts over time and correlated these with the classification of patient mutation subtypes. Most participants experienced age-related declines in hematologic parameters beginning in early childhood. Platelets underwent the earliest decline, reaching platelet count below 50K/μl at a median age of 8.2 years. The erythrocyte lineage was the most stable with hemoglobin below 8 g/dl identified at a median age of 10.7 years. Androgen therapy delayed the blood count decline. The presence of at least one predicted hypomorphic pathogenic variant in the *FANCA* gene significantly slowed the progression of the hematologic abnormalities. This study sheds light on the importance of mutation type in predicting the severity of hematological manifestations in FA. Furthermore, it serves as a historical comparative cohort for emerging therapies aimed at altering hematological disease progression in *FANCA-*deficient FA patients.

**Key Points:**

- Thrombocytopenia and neutropenia are the earliest and most reliable indicators of hematologic decline in *FANCA-*deficient patients
- Presence of two *FANCA* variants with predicted complete loss of function leads to earlier onset and faster progression of disease

## Introduction

Fanconi anemia (FA) is an inherited DNA repair disorder ^1^. Its clinical characteristics include congenital anomalies, childhood-onset progressive bone marrow failure (BMF) and lifelong increased cancer risk ^2,3^. Approximately eighty percent of FA patients have at least 1 congenital abnormality^4–7^. Premalignant and malignant conditions associated with FA include myelodysplastic syndrome (MDS), acute myeloid leukemia (AML) and solid malignancies, most frequently squamous cell carcinoma (SCC) of the head and neck ^8–10^.

The clinical diagnosis of FA is typically established by increased cellular sensitivity and chromosomal breakage of lymphocytes and/or skin fibroblasts in response to treatment with diepoxybutane (DEB) or mitomycin C (MMC)^11^.

There are 23 currently recognized FA-causing genes, *FANCA* through *FANCX* ^3,12,13^. Of these, all are inherited in an autosomal recessive pattern, except for *FANCB*, which is X-linked recessive^14^, and *FANCR*, which is autosomal dominant and *de novo*^15–17^. Onset and severity of hematologic abnormalities in patients with *FANCB* have been found to correlate with the level of residual FANCD2 monoubiquitination^18^, while patients with *FANCM, FANCR, FANCO and FANCU* pathogenic variants (which we may refer to as variants or mutations throughout the text) have not developed bone marrow failure unless challenged with chemotherapy treatment^15–17,19–22^. *FANCA* is the most commonly mutated gene in FA, accounting for sixty to seventy percent of all affected patients depending on a cohort ^18,23,24^. The FANCA protein is a component of the FA core complex which is necessary for the ubiquitination of FANCI and FANCD2, proteins which then initiate the downstream repair process^2^. The spectrum of *FANCA* pathogenic variants is heterogenous and includes single nucleotide variants, small insertions and deletions (indels), truncations, and large deletions^25^. The majority of these variants are unique to the individuals carrying them, with the exception of a few, including known founder mutations ^26–29^. A previous evaluation of FA patients from 1989-1999 involved data analysis from multiple complementation groups and found that patients with null *FANCA* mutations exhibited earlier onset of hematologic dysfunction, shorter survival after diagnosis and higher incidence of MDS and AML compared to those with mutations that produced an altered protein product, presumably with some level of residual functional protein^30^. The complexity of genotype-phenotype correlation is further increased by the phenomenon of somatic mosaicism, which has been observed in several subtypes of FA, including those caused by *FANCA* variants. Mosaicism can arise through various mechanisms, including reversion or other compensatory mutations in hematopoietic stem cells, leading to the coexistence of both affected and unaffected cell populations within the same individual. This often results in a milder clinical phenotype^31–34^.

The average age of onset of hematologic abnormalities in patients with FA is approximately 7.6 years^30^, but the course of hematologic decline varies widely. Following onset of hematologic abnormalities, adjunct therapies such as androgens have been utilized to provide interval improvements in blood counts while awaiting definitive therapy, such as allogeneic hematopoietic stem cell transplant (HSCT)^35^. Indications for HSCT in FA include severe BMF nearing transfusion need or reduced neutrophil counts increasing the risk of infection, or development of MDS or AML. Due to their underlying genetic defect and inherent sensitivity to genotoxic agents, patients with FA historically have had higher rates of complications associated with HSCT. Over the past several decades; however, modifications to pre-HSCT conditioning protocols for patients with FA have reduced toxicity and significantly improved outcomes^36–40^.

Despite these modifications to therapy and improvements in survival, FA patients treated by allogeneic HSCT are at risk of graft vs. host disease (GVHD) in the absence of T cell depletion and an increased risk of solid tumors compared to FA patients not requiring transplant^41,42^. This has led to a search for alternative approaches for treating FA-related BMF, for example gene therapy approaches^43^. As consideration of gene therapy requires the availability of adequate numbers of HSCs for genetic modification, there is a critical need to determine the optimal timing for such an intervention in patients with FA to achieve maximal yield of HSCs. We would also be helped by the knowledge of when FA patients may need transplant.

Here, we report detailed analysis of *FANCA* pathogenic variants and their impact on the trajectory of blood count decline. We found that thrombocytopenia is the earliest indicator of hematologic decline in most patients with FA, that androgens have been effective in rescuing observed decreases in cell counts prior to the need for transplant and that the presence of at least one variant predicted to have residual function often leads to slower decline in blood counts.

## Methods

### Study Participants

Most of the participants in this study were enrolled in the International Fanconi Anemia Registry (IFAR) after provision of written informed consent and assent, where applicable. The IFAR database was filtered for all participants with pathogenic variants in *FANCA* and for those patients diagnosed after 1995, were diagnosed prior to age 18 years, with at least 2 pre-transplant complete blood count values a minimum of 30 days apart (total *n* = 125). Participants with known blood mosaicism were excluded. Diagnosis of mosaicism was based on clinical breakage test concluding presence of mosaicism or molecular studies showing a mechanism of reversion (Chandrasekharappa and Smogorzewska labs, unpublished). Exclusion of patients diagnosed after age 18 was performed based on the recognition that many of the known patients diagnosed in adulthood have blood mosaicism or express hypomorphic alleles (^30,34,44–47^, and Chandrasekharappa and Smogorzewska labs, unpublished). To increase the number of individuals, data from an additional 14 patients not enrolled in IFAR, but meeting the criteria of the study were obtained from Cincinnati Children’s Hospital Medical Center (CCHMC). Data was extracted regarding congenital anomalies, blood count parameters, medical interventions (inclusive of medications, transfusions, surgeries and HSCT) and clinical outcomes (including occurrence of cancers). Additional information was obtained from participants, their families, and their treating physicians to supplement existing IFAR data. End of follow-up was January 1^st^ 2021. Institutional Review Board (IRB) of the Rockefeller University gave ethical approval for this work. IRB of the Cincinnati Children’s Hospital Medical Center gave ethical approval for the work involving non-IFAR participants.

### Summary of collected data

Records from 139 patients (125 patients from the IFAR and 14 patients from CCHMC, but not in IFAR) with a total of 1806 visits were extracted. There were 1795 entries for platelet count, white blood count (WBC), and hemoglobin (Hb). There were 1588 entries for absolute neutrophil count (ANC), and 1491 entries for mean corpuscular volume (MCV). 1356 entries corresponded to data from individuals not on an androgen, 302 entries from individuals on androgens, and 148 entries with unknown androgen treatment status.

### *FANCA* Mutation Analysis

*FANCA* variants were categorized into missense, nonsense, splicing, in-frame deletions and large deletions^25^. SpliceAI^48^ was used to predict variants affecting splicing. For comparisons shown in Figure 4 and Table 3, large deletions and nonsense variants were considered to be loss of function alleles with the remainder of the variants predicted to retain residual function (i.e., considered to be hypomorphic alleles).

### Statistical Analysis

A linear mixed-effects model was used to compare the change in blood count parameters over time by androgen therapy status. The model included age, androgen therapy status as a time-varying covariate, and interaction between age and androgen therapy status, with random intercept and slope for each participant (Proc Mixed, SAS Studio 3.8). Patient visits with unknown androgen therapy status (148 visits from 16 patients) were excluded from this analysis. Patients who were noted to be receiving GCSF (n = 19) were excluded from all analyses of total WBC and ANC due to extreme variations in these counts. Log transformations of blood counts were performed prior to all analyses to reduce heterogeneity of variance due to variability in the spread of blood counts over the age range, and to meet the normality assumption. Blood count data were visualized as spaghetti plots over time for all participants with available data (n = 123 for platelets/hemoglobin/mean corpuscular volume (MCV), n = 104 for WBC/ANC excluding GCSF), with each line representing an individual participant. Estimated slopes and loess smoother curves were plotted. Linear mixed-effects model with random intercept and slope was also used for comparing change in blood counts over time by *FANCA* mutation status after excluding patient visits with androgen therapy. Information on transfusion of blood products was incomplete. In cases in which transfusion of packed red blood cells and platelets was noted, the pretransfusion value was used if available. We noted that transfusions had only mild, transient effects on the post-transfusion values, without visible effect on the overall slope, therefore participants receiving transfusions were included in all analyses.

## Results

### Participant demographics, clinical phenotypes and outcomes

Characteristics of the 139 study participants are listed in **Table 1**. The mean age at last follow-up, including the deceased participants, was 19.3 (range of 4.2-41.0) years and for living participants was 20.7 years (range 4.2-41.0 years). The cohort was comprised of a majority white, non-Hispanic population. The male: female ratio was 1.1:1. The mean age at diagnosis of FA was 6.3 years (range of birth to 18 years). One hundred thirty-one participants (94.2%) had at least one congenital abnormality documented and 67 (48.2%) had 3 or more such anomalies (**Table 2**). The full spectrum of phenotypes is provided in **Supplemental Table 1**. A majority (78.4%) of participants underwent HSCT, with the mean age at HSCT of 10.4 years (range of 3.3 years to 27.9 years). Thirteen (9.4%) participants had history of MDS, six (4.3%) developed AML (n=6; 4.3%), 21 (15.1%) developed solid tumors. One patient (0.7%) developed Post-Transplant Lymphoproliferative Disorder (PTLD) (n=1; 0.7%). At time of data analysis, 71.9% (n = 100) of participants were alive, 18.0% (n = 25) had died, and 10.1% (n = 14) had unknown vital status. The mean age of death was 16.4 years (range of 4.7 years to 32.0 years). Causes of death were transplant-related complications (n = 12), AML (n = 1), squamous cell carcinoma (n = 5), osteosarcoma (n=1), liver cancer (n=1), and other (aspiration pneumonia, sepsis, multiorgan failure, and two unknown causes).

**Table 1:** Subject Demographics.

|  | **n** | **%** |
| --- | --- | --- |
| **Gender** |  |  |
| Male | 72 | 51.8 |
| Female | 67 | 48.2 |
| **Race** |  |  |
| White | 109 | 78.4 |
| Asian | 6 | 4.3 |
| Black/African American | 5 | 3.6 |
| Other | 11 | 7.9 |
| Unknown | 8 | 5.8 |
| **Ethnicity** |  |  |
| Non-Hispanic | 119 | 85.6 |
| Hispanic | 15 | 10.8 |
| Unknown | 5 | 3.6 |
| **HSCT** |  |  |
| Yes | 109 | 78.4 |
| No | 24 | 17.3 |
| Unknown | 6 | 4.3 |

**Table 2.** Congenital abnormalities in individuals with *FANCA* mutations. The value excludes short stature. Mean number of abnormalities per patient = 2.7

|  | **n** | **%** |
| --- | --- | --- |
| Skeletal | 97 | 69.8 |
| Skin | 79 | 56.8 |
| Cardiac | 21 | 15.1 |
| Gastrointestinal | 23 | 16.6 |
| Genitourinary | 50 | 36.0 |
| Microcephaly | 26 | 18.7 |
| Other CNS | 3 | 2.2 |
| Eye | 33 | 23.7 |
| Ear | 40 | 28.8 |

**Table 3a.** Significance of differences in blood count decline over time by type of *FANCA* mutations in the cohort of 139 individuals with Fanconi anemia. Comparison 1 was performed between individuals with at least one missense variant and individuals with combination of nonsense, frameshift, in-frame, splicing, and large deletions. Comparison 2 was performed between individuals with at least one missense variant and individuals with combination of nonsense, frameshift, and large deletions. Comparison 3 was performed between individuals with at least one missense, in-frame, or splicing variant and individuals with combination of nonsense, frameshift, and large deletions. Comparison 4 was performed between individuals with both variants of missense, in-frame, or splicing type and individuals with combination of nonsense, frameshift, and large deletions only. Comparison 5 was performed between individuals with both missense variants and individuals with combination of nonsense, frameshift, and large deletions only. Sample size was much smaller in comparison 5. Age-by-mutation group interaction p-values are shown in the table for each comparison. Data for comparison 3 which was testing our hypothesis is shown in **Figure 4**.

|  | Comparison 1 | Comparison 2 | Comparison 3 | Comparison 4 | Comparison 5 |
| --- | --- | --- | --- | --- | --- |
| Log PLT | **0.0022** | **0.0001\*** | **<0.0001\*** | **0.0059** | 0.2492 |
| Log WBC | **0.0002\*** | **0.0001\*** | **<0.0001\*** | **0.0011\*** | 0.8220 |
| Log Hb | **0.0231** | **0.0273** | 0.0956 | 0.1121 | 0.9722 |
| Log ANC | **0.0038** | **0.0004\*** | **0.0006\*** | **<0.0001\*** | 0.2512 |
| Log MCV | 0.6311 | 0.7708 | 0.9924 | 0.8675 | 0.7791 |

p values < 0.05 are in bold. p-values that meet the Bonferroni adjusted alpha level of 0.05/25 tests <0.002 are indicated with the asterisk (\*).

**Table 3b.** Comparison 3 from Table 3a further broken down by androgen status. 3a: excludes androgen therapy visits (comparison 3 from above), 3b: includes pre-androgen therapy visits only, 3c: excludes any subjects given androgen therapy

|  | Comparison 3a | Comparison 3b | Comparison 3c |
| --- | --- | --- | --- |
| Log PLT | <0.0001 | <0.0001 | <0.0001 |
| Log WBC | <0.0001 | 0.0001 | <0.0001 |
| Log Hb | 0.0956 | 0.0729 | 0.0326 |
| Log ANC | 0.0006 | 0.0016 | 0.0007 |
| Log MCV | 0.9924 | 0.9681 | 0.9181 |

### Onset of hematologic abnormalities

Pre-transplant blood counts were plotted over time for all participants (**Fig 1**). Median number of CBCs per patient was 8 with a median interval time between first and last CBCs of 3 years. Transfusions of packed red blood cells and platelets were noted when these data were available. Note was also made of concurrent therapies including androgens, corticosteroids, and GCSF (see sample detailed individual participant plots in **Supplemental Figure 1**). Blood product transfusion was noted to have only transient effects on post-transfusion values and minimal effect on overall count trends, therefore participants receiving transfusions were included in all analyses. Participants who received GCSF (n = 19) were noted to have extreme variations in WBC-related values and were therefore excluded from all analyses involving WBC or ANC. These participants are plotted separately and shown in **Supplemental Figure 2**.

**Figure 1.**
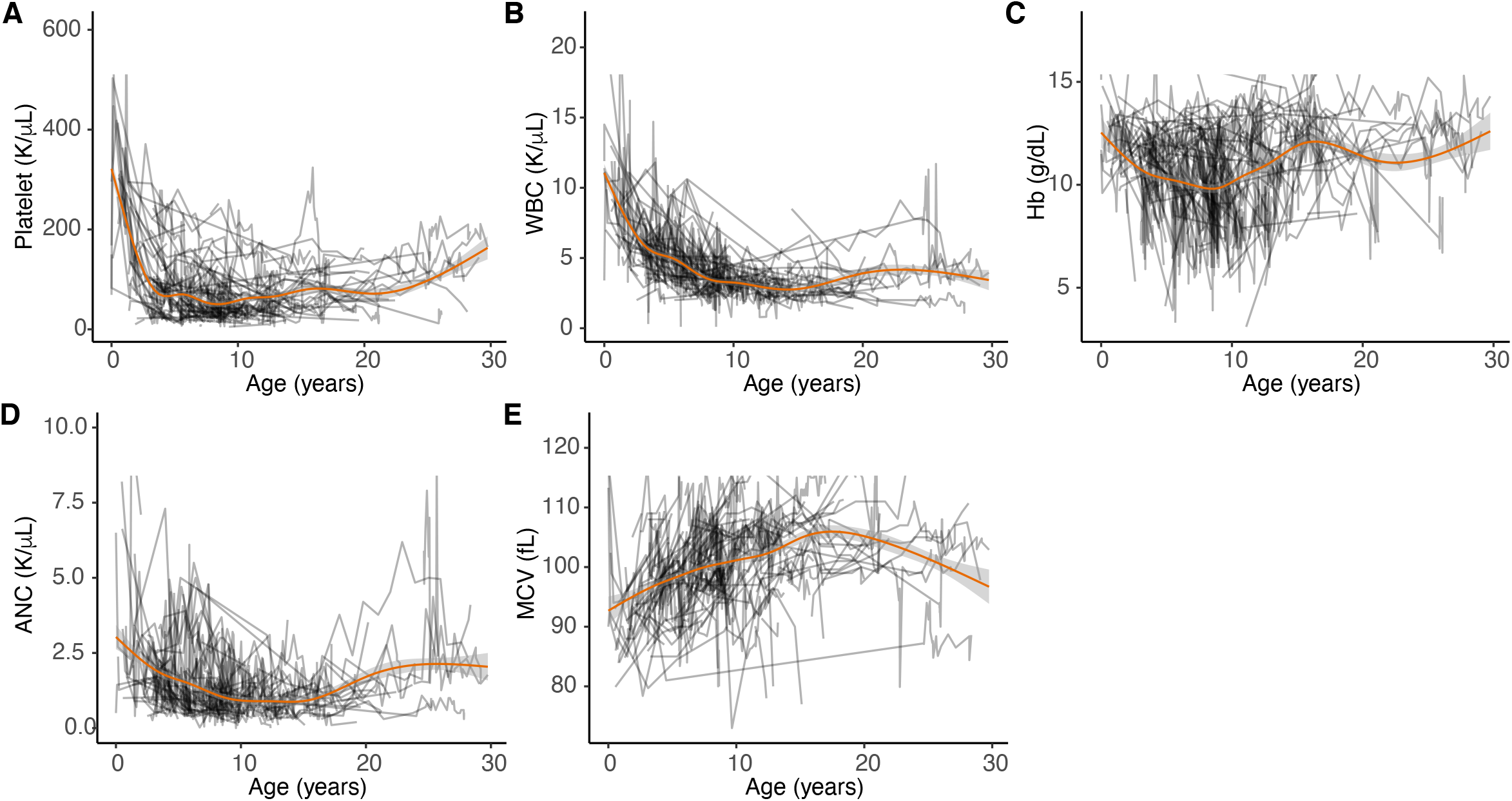
Kinetics of blood count decline in the cohort of 139 individuals with Fanconi anemia caused by FANCA pathogenic variants. Spaghetti plots over time (in years) with each line representing an individual participant for the indicated counts: A. Platelet count B. White Blood Count (WBC) C. Hemoglobin D. Absolute Neutrophil Count (ANC) E. Mean Corpuscular Volume (MCV). n = 123 for platelets/hemoglobin/mean corpuscular volume; n = 104 for White Blood Count /Absolute Neutrophil Count excluding those treated with GCSF. Estimated Loess smoother curves are shown in orange.

Overall (n=139), a one-year increase in age was associated with a 12.2% decrease in platelets, 7.9% decrease in WBC (n = 120 excluding GCSF treated patients), 2.5% decrease in Hb, 7.6% decrease in ANC (n = 120 excluding GCSF treated patients) and 0.9% increase in MCV. The decreases reflect the overall average trend observed in the participants, with the steepest decrease occurring in the first decade of life.

Kaplan-Meier survival analysis was performed to calculate the median age at which each cell lineage experienced clinically relevant decline. Thresholds were set as follows for moderate and severe cytopenia respectively. For neutropenia: ANC <1.0K/μl and <0.5K/μl. thrombocytopenia: platelets <50K/μl and <25K/μl; anemia: hemoglobin <8g/dl and <6g/dl. 130 (93.5%) patients experienced at least 1 moderate grade cytopenia. Neutropenia was the most common hematologic abnormality, with 81% of participants experiencing moderate grade, and 50% of participants experiencing severe grade with median ages at onset of 8.4 years and 11.9 years respectively (**Fig 2A and B**). Thrombocytopenia was the second most common hematologic abnormality, with 78% of participants experiencing moderate grade and 47% of participants experiencing severe grade, with median ages at onset of 8.2 years and 11.1 years respectively (**Fig 2C and D**). Hemoglobin was the least severely affected parameter with 50% of participants experiencing moderate anemia at a median age of 10.7 years (**Fig 2E**). 14% of participants developed severe anemia; however, a median age was not reached, since most patients underwent HSCT before reaching this criterion (**Fig 2F**).

**Figure 2.**
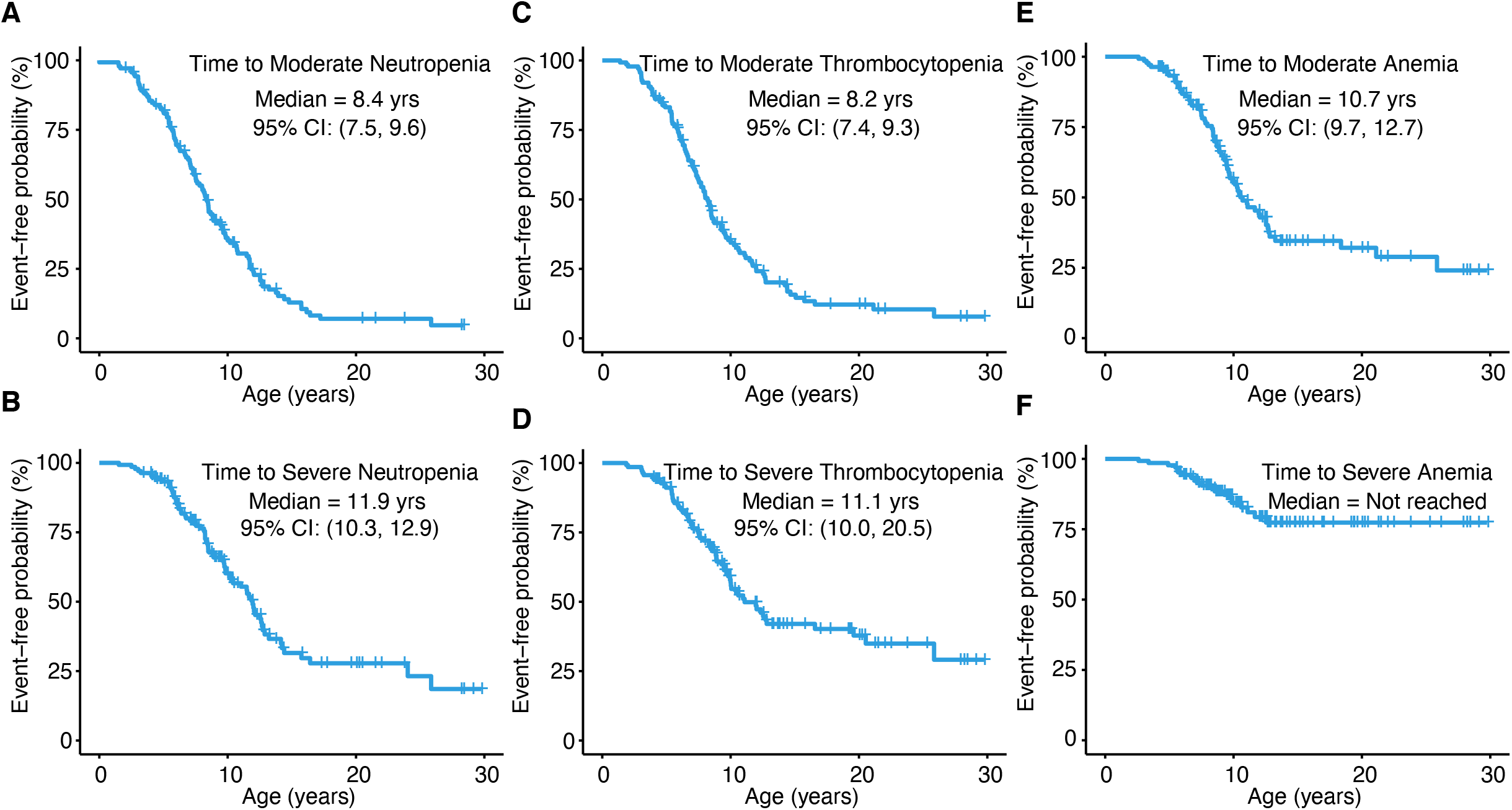
Kaplan Meier analysis of time to neutropenia, thrombocytopenia, and anemia development in individuals with Fanconi anemia caused by FANCA pathogenic variants. A. Time to moderate neutropenia; ANC <1,000/μl 112 events (81%), 27 censored (19%) B. Time to severe neutropenia; ANC <500/ μl: 70 events (50%), 69 censored (50%) C. Time to moderate thrombocytopenia; platelets <50,000/ μl: 108 events (78%), 31 censored (22%) D. Time to severe thrombocytopenia; Platelets <25,000/μl: 66 events (47%), 73 censored (53%) E. Time to moderate anemia; Hemoglobin <8g/dL: 70 events (50%), 69 censored (50%) F. Time to severe anemia; Hemoglobin <6g/dL: 20 events (14%), 119 censored (86%).

### Effect of androgen use on hematopoietic failure

We documented the presence or absence of androgen therapy for every patient visit collected and plotted CBC values on the original scale (**Supplemental Figure 3**). After log transformation of blood count parameters for analysis (see methods section), we compared the slopes of cell counts over time by androgen therapy status. Androgen use was found to be a significant predictor of change in cell counts when included as a time varying covariate. We observed significant age-by-androgen therapy interaction effects in all the blood counts analyzed (interaction p-values ≤ 0.001). When patients were not on androgen therapy, a one-year increase in age was associated with a 15.8% decrease in platelets, 8.9% decrease in WBC, 3.6% decrease in Hb, 9.0% decrease in ANC and 1.1% increase in MCV. In contrast, when patients were on androgens, a one-year increase in age was associated with a 0.1% decrease in platelets, 2.3% decrease in WBC, 0.9% increase in Hb, 2.7% increase in ANC, and 0.3% increase in MCV (**Fig 3 A-E**).

**Figure 3.**
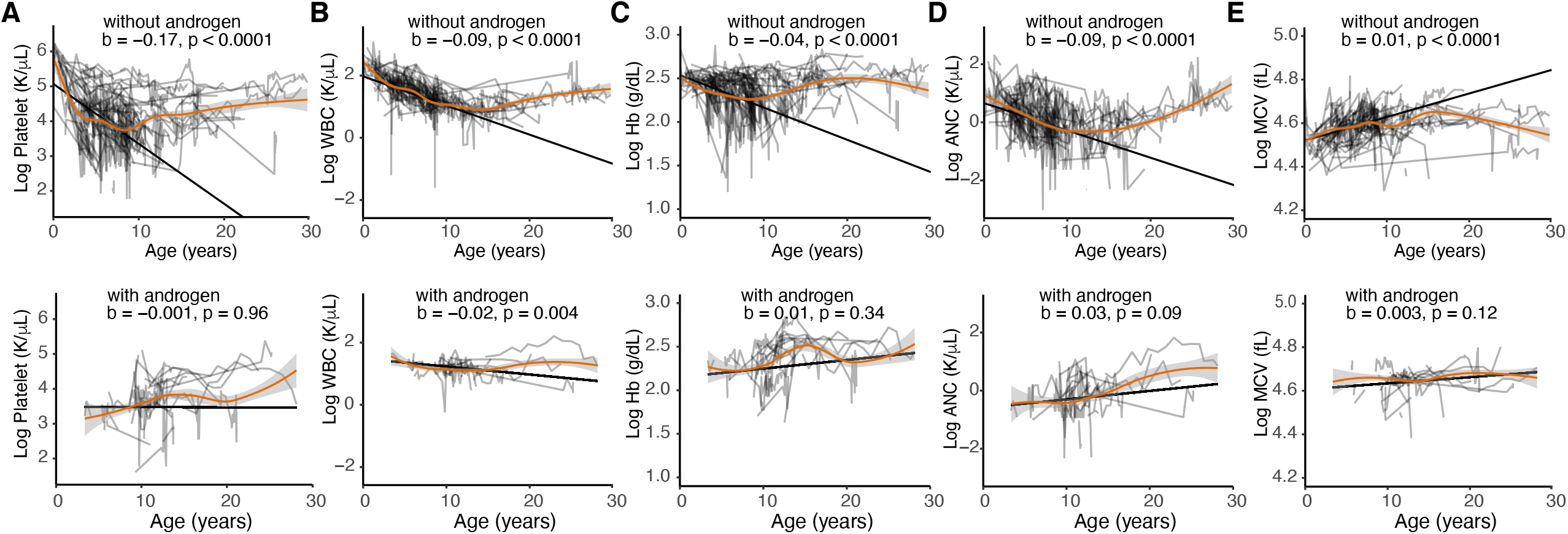
Effects of Androgen on the kinetics of blood count decline in the cohort of 139 individuals with Fanconi anemia caused by FANCA pathogenic variants. A. Platelet count B. White Blood Count (WBC) C. Hemoglobin D. Absolute Neutrophil Count (ANC) E. Mean Corpuscular Volume (MCV). Spaghetti plots over time (in years) with each line representing an individual participant. Counts obtained while an individual was on Androgen are shown in the bottom row. Counts with unknown androgen therapy status were excluded. White Blood Count /Absolute Neutrophil Count for patients treated with GCSF were also excluded. Estimated slopes and loess smoother curves are shown.

### *FANCA* spectrum of pathogenic variants varies widely and correlates with severity of cytopenias

The spectrum of *FANCA* pathogenic variants observed was highly heterogeneous. Of the mutations, 48.2% were substitutions, 43.9% were deletions, 5.0% were duplications and 2.9% were unknown. The expected effects of these variants included 18.0% missense, 13.7% nonsense, 19.8% frameshift, 8.3% in-frame deletions, 16.9% splice site, 19.0% large deletion, and 4.3% were undetermined. The complete listing of all patient mutations and predicted outcomes is provided in Supplemental Table 1.

We hypothesized that participants with both *FANCA* variants coded as either nonsense, frameshift or large deletion, would have a more severe phenotype due to the expected complete lack of FANCA protein function. Participants with at least one mutation coded as missense, in-frame or splicing, would then display a milder clinical phenotype due to predicted residual FANCA levels and activity. To test our hypothesis, we investigated the age of hematologic onset in patients with FA carrying two predicted null variants compared to those carrying at least one mutation predicted to produce protein with some residual function. The slopes for hematopoietic lineage decline between groups were significantly different with age-by-mutation group interaction p-values < 0.001 for platelets, WBC and ANC (**Table 3, comparison 3**). For patients with predicted null mutations, a one-year increase in age was associated with a 24.5% decrease in platelet count, 15.1% decrease in WBC count and 15.4% decrease in ANC. In contrast, those patients with at least one variant predicted to result in one partially functioning protein had a 12.1% decrease in platelets, 6.9% decrease in WBC count and 6.3% decrease in ANC (**Fig 4**). For completeness of the analysis, we performed additional comparisons between expected variant effect and the blood count decline (**Table 3**) with presence of a missense variant being a strong predictor of better outcome.

**Figure 4.**
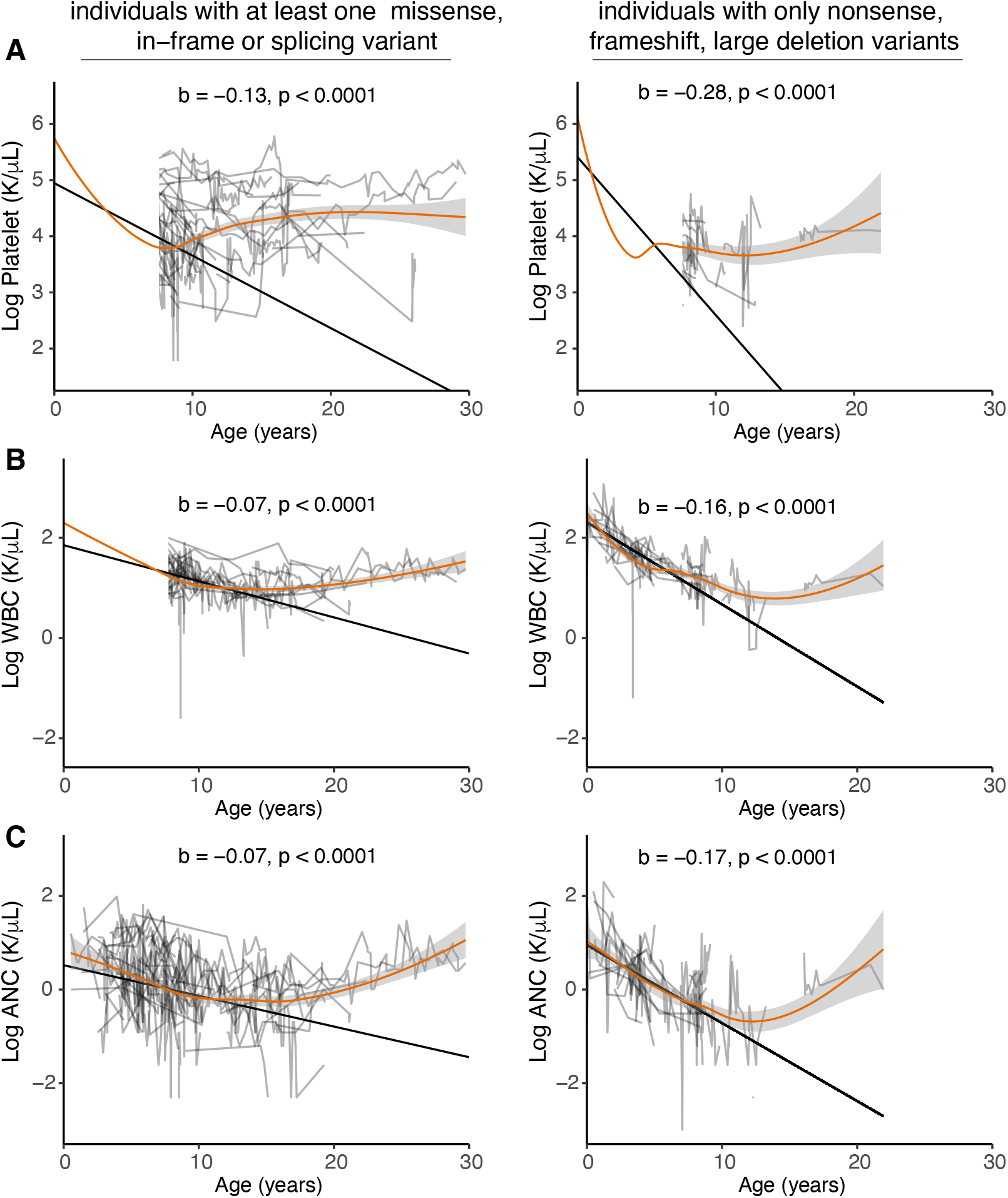
Effects of FANCA mutation type on the kinetics of blood count decline in the cohort of 139 individuals with Fanconi anemia. Spaghetti plots over time (in years) with each line representing an individual participant for the indicated counts: A. Platelet count B. White Blood Count (WBC) C. Absolute Neutrophil Count (ANC). Left column depicts individuals with at least one missense, in-frame, or splicing variant and the right column depicts individuals with combination of nonsense, frameshift, and large deletions. Also see Table 3 for a summary of p-values for other group comparisons.

## Discussion

This study examined the natural history of hematologic progression in a contemporary cohort of patients carrying *FANCA* pathogenic variants. We have been able to stratify the hematologic decline in our cohort according to cell lineage, finding that platelets and neutrophils are the most sensitive indicators of hematologic onset. We have also confirmed that androgens are effective in reversing the trend of hematologic decline and can be utilized as a bridge to a possible HSCT ^35^, notwithstanding the known risks associated with its usage including liver toxicity and increased mortality during subsequent HSCT ^49^. The majority of our cohort (n=109, 78.4%) underwent a bone marrow transplantation. Of those, 12 patients (11%) had died from the bone marrow transplantation complications. Since our cohort drew from patients diagnosed since 1995, this high number of BMT complications may not reflect current BMT outcomes and readers are directed to recent studies describing outcomes in FA patients^36–40,50^.

High variability of the disease presentation and course has been seen in patients with *FANCA* mutations ^4^. One of the reasons might be that the majority of *FANCA* pathogenic variants are unique to each family. By closely examining mutational status and predicted protein outcomes, we identified groups at either end of the spectrum of clinical variability. Patients with two predicted *FANCA* null alleles were found to have earlier hematologic onset and more severe outcomes, which is consistent with a prior study describing a smaller number of *FANCA* patients^30^.

Our cohort also included a few patients with no significant hematologic abnormalities even into the third decade of life. Although our cohort excluded subjects that were diagnosed as mosaics through clinical chromosome breakage testing or mosaicism proven by molecular studies (Chandrasekharappa and Smogorzewska labs, unpublished), it is possible that these patients developed mosaicism after their breakage or molecular studies were completed, resulting in missed mosaicism diagnosis. Alternatively, given that the majority of these individuals had point mutations in *FANCA*, this could indicate they carried hypomorphic mutations with significant residual FANCA function that did not lead to blood count declines. As we and others have shown, FANCA with pathogenic amino acid substitution is most often localized to the cytoplasm ^25,51–53^, and upon envelope breakdown some FANCA, may localize to the DNA where it could function.

Limitations of our study include its retrospective nature, and its dependence on the quality of available medical records. We note that the documentation of blood product transfusion and regimens of adjunct medications used in FA treatment were of varying quality between individual medical records. Despite this, even with the relatively small proportion of our cohort with documented androgen use, we were able to establish significant differences in clinical parameters over time by androgen therapy status. In comparing our records with documented transfusions that were reflected in the blood counts, we noted that the impact of these transfusions was transient to the overall trend of declining blood counts over time and were unlikely to have had significant impact on the time to HSCT (See **Supplemental Fig 1A and B**). This is underscored by the fact that current clinical practice is restrictive regarding blood product transfusions to prevent adverse impact from alloimmunization.

The length of longitudinal follow-up required for evaluation in our cohort may have biased the study towards patients whose clinical status on presentation was manageable without HSCT for a period of at least one year. Subjects with severe disease or precipitous decline in counts prior to, or immediately after diagnosis, who proceeded to HSCT within months, would have been excluded from our cohort based on absence of longitudinal blood count data. Notably, the fact that our records were almost exclusively obtained from HSCT referral centers may also have contributed to skewing of the median age of hematologic onset. Since most of our data was after a referral had been triggered clinically, it is possible that the actual time of hematologic onset was even earlier than our results demonstrate.

Despite these limitations, our study represents the largest cohort of patients with *FANCA* pathogenic variants that has been evaluated with this longitudinal breadth and degree of data granularity. A prospective study in the context of the IFAR would not have been feasible as patients were treated in a variety of healthcare settings nationally and internationally, prior to their referral to a major US center specializing in HSCT for FA. Our results provide a contextual view of the overall clinical progression of patients with *FANCA* mutations leading up to HSCT. This represents useful baseline data upon which to evaluate the efficacy of clinical trials of gene and any other therapy. It also provides useful information about the typical age at which hematologic onset is expected to occur, which is a key parameter in every intervention, and more so in gene therapy approaches, as patients’ HSCs must be harvested well before this deterioration begins for optimal cell yields.

Future work should include prospective longitudinal studies of this patient population, with subsequent focus on patient subgroups with extremes in age of hematologic onset. Clarifying the extent to which hypomorphic mutations or mosaicism plays a role in these clinical outcomes could also be informative as we increase our understating of the proportion of functional FANCA protein that is necessary to obtain improvement in clinical outcomes.

## Data Availability

All data produced in the present work are contained in the manuscript

## Acknowledgements

We are indebted to the generosity of the FA patients and families, as well as their treating physicians, that participated in this study. This work was supported by grants from the National Institutes of Health (NIH)); National Cancer Institute (R01 CA204127) (AS); National Center for Advancing Translational Sciences (KL2TR001865) (RRM), (UL1 TR001866) (RRM, CSJ, TB, and AS) and Stavros Niarchos Foundation (TB). Rocket Pharmaceuticals provided partial salary support of AS and full salary support of AG and AMG and provided administrative support for record procurement over the duration of the study. AS was a Howard Hughes Medical Institute Faculty Scholar. FXD and SCC acknowledge support from the Intramural Research Program of the NIH National Human Genome Research Institute.

## Author Contributions

RRM and AS conceived and designed this study. RRM, AS, AR, AMG, TB and YCL aggregated cohort data from the IFAR. SCC, FXD, DCK, SS and FPL performed molecular and sequencing analysis of patient mutations. FB, MIC, JK, SMD, PAM, JO, MLM, JEW and RT provided clinical information for subjects. CSJ performed biostatistical analysis of the data. ADA directed clinical, complementation group testing and sequencing analysis in the subjects before 2009. RRM, TB and AS wrote the manuscript with essential input from other authors. AS acquired funding and supervised.

## Conflict of Interest Disclosure

AR and AMG obtained salary support and AS, partial salary support from Rocket Pharmaceuticals for this study. AS and JW are advisors to Rocket Pharmaceuticals. The remaining authors declare no competing financial interests.

## Notes

### Competing Interest Statement

AR and AMG obtained salary support and AS, partial salary support from Rocket Pharmaceuticals for this study. AS and JW were advisors to Rocket Pharmaceuticals. The remaining authors declare no competing financial interests.

### Author Declarations

Institutional Review Board (IRB) of the Rockefeller University gave ethical approval for this work. IRB of the Cincinnati Childrens Hospital Medical Center gave ethical approval for the work involving non-IFAR participants.

## References

1. Taylor AMR, Rothblum-Oviatt C, Ellis NA, et al. Chromosome instability syndromes. Nat Rev Dis Primers. Sep 19 2019;5(1):64. doi:10.1038/s41572-019-0113-0

2. Kottemann MC, Smogorzewska A. Fanconi anaemia and the repair of Watson and Crick DNA crosslinks. Nature. Jan 17 2013;493(7432):356–63. doi:10.1038/nature11863

3. Niraj J, Farkkila A, D’Andrea AD. The Fanconi Anemia Pathway in Cancer. Annu Rev Cancer Biol. Mar 2019;3:457–478. doi:10.1146/annurev-cancerbio-030617-050422

4. Fiesco-Roa MO, Giri N, McReynolds LJ, Best AF, Alter BP. Genotype-phenotype associations in Fanconi anemia: A literature review. Blood Rev. Sep 2019;37:100589. doi:10.1016/j.blre.2019.100589

5. Alter BP, Giri N. Thinking of VACTERL-H? Rule out Fanconi Anemia according to PHENOS. Am J Med Genet A. Jun 2016;170(6):1520–4. doi:10.1002/ajmg.a.37637

6. Giampietro PF, Adler-Brecher B, Verlander PC, Pavlakis SG, Davis JG, Auerbach AD. The need for more accurate and timely diagnosis in Fanconi anemia: a report from the International Fanconi Anemia Registry. Pediatrics. Jun 1993;91(6):1116–20.

7. Alter BP, Rosenberg PS. VACTERL-H Association and Fanconi Anemia. Mol Syndromol. Feb 2013;4(1-2):87–93. doi:10.1159/000346035

8. Alter BP, Giri N, Savage SA, Rosenberg PS. Cancer in the National Cancer Institute inherited bone marrow failure syndrome cohort after fifteen years of follow-up. Haematologica. Jan 2018;103(1):30–39. doi:10.3324/haematol.2017.178111

9. Kutler DI, Auerbach AD, Satagopan J, et al. High incidence of head and neck squamous cell carcinoma in patients with Fanconi anemia. Arch Otolaryngol Head Neck Surg. Jan 2003;129(1):106–12. doi:10.1001/archotol.129.1.106

10. Webster ALH, Sanders MA, Patel K, et al. Genomic signature of Fanconi anaemia DNA repair pathway deficiency in cancer. Nature. Dec 2022;612(7940):495–502. doi:10.1038/s41586-022-05253-4

11. Auerbach AD. Fanconi anemia and its diagnosis. Mutat Res. Jul 31 2009;668(1-2):4–10. doi:10.1016/j.mrfmmm.2009.01.013

12. Harrison BA, Mizrahi-Powell E, Pappas J, et al. Deficiency of the Fanconi anemia core complex protein FAAP100 results in severe Fanconi anemia. J Clin Invest. Apr 17 2025; doi:10.1172/JCI185126

13. Kuehl J, Xue Y, Yuan F, et al. Genetic inactivation of FAAP100 causes Fanconi anemia due to disruption of the monoubiquitin ligase core complex. J Clin Invest. Apr 15 2025; doi:10.1172/JCI187323

14. Meetei AR, Levitus M, Xue Y, et al. X-linked inheritance of Fanconi anemia complementation group B. Nat Genet. Nov 2004;36(11):1219–24. doi:10.1038/ng1458

15. Ameziane N, May P, Haitjema A, et al. A novel Fanconi anaemia subtype associated with a dominant-negative mutation in RAD51. Nat Commun. Dec 18 2015;6:8829. doi:10.1038/ncomms9829

16. Wang AT, Kim T, Wagner JE, et al. A Dominant Mutation in Human RAD51 Reveals Its Function in DNA Interstrand Crosslink Repair Independent of Homologous Recombination. Mol Cell. Aug 6 2015;59(3):478–90. doi:10.1016/j.molcel.2015.07.009

17. Takenaka S, Kuroda Y, Ohta S, et al. A Japanese patient with RAD51-associated Fanconi anemia. Am J Med Genet A. Jun 2019;179(6):900–902. doi:10.1002/ajmg.a.61130

18. Jung M, Ramanagoudr-Bhojappa R, van Twest S, et al. Association of clinical severity with FANCB variant type in Fanconi anemia. Blood. Apr 30 2020;135(18):1588–1602. doi:10.1182/blood.2019003249

19. Bogliolo M, Bluteau D, Lespinasse J, et al. Biallelic truncating FANCM mutations cause early-onset cancer but not Fanconi anemia. Genet Med. Apr 2018;20(4):458–463. doi:10.1038/gim.2017.124

20. Catucci I, Osorio A, Arver B, et al. Individuals with FANCM biallelic mutations do not develop Fanconi anemia, but show risk for breast cancer, chemotherapy toxicity and may display chromosome fragility. Genet Med. Apr 2018;20(4):452–457. doi:10.1038/gim.2017.123

21. Park JY, Virts EL, Jankowska A, et al. Complementation of hypersensitivity to DNA interstrand crosslinking agents demonstrates that XRCC2 is a Fanconi anaemia gene. J Med Genet. Oct 2016;53(10):672–680. doi:10.1136/jmedgenet-2016-103847

22. Vaz F, Hanenberg H, Schuster B, et al. Mutation of the RAD51C gene in a Fanconi anemia-like disorder. Nat Genet. May 2010;42(5):406–9. doi:10.1038/ng.570

23. Mehta PA, Ebens C. Fanconi Anemia. In: Adam MP, Mirzaa GM, Pagon RA, et al, eds. GeneReviews((R)). 1993.

24. Gille JJ, Floor K, Kerkhoven L, Ameziane N, Joenje H, de Winter JP. Diagnosis of Fanconi Anemia: Mutation Analysis by Multiplex Ligation-Dependent Probe Amplification and PCR-Based Sanger Sequencing. Anemia. 2012;2012:603253. doi:10.1155/2012/603253

25. Kimble DC, Lach FP, Gregg SQ, et al. A comprehensive approach to identification of pathogenic FANCA variants in Fanconi anemia patients and their families. Hum Mutat. Feb 2018;39(2):237–254. doi:10.1002/humu.23366

26. Tipping AJ, Pearson T, Morgan NV, et al. Molecular and genealogical evidence for a founder effect in Fanconi anemia families of the Afrikaner population of South Africa. Proc Natl Acad Sci U S A. May 8 2001;98(10):5734–9. doi:10.1073/pnas.091402398

27. Callen E, Casado JA, Tischkowitz MD, et al. A common founder mutation in FANCA underlies the world’s highest prevalence of Fanconi anemia in Gypsy families from Spain. Blood. Mar 1 2005;105(5):1946–9. doi:10.1182/blood-2004-07-2588

28. Amouri A, Talmoudi F, Messaoud O, et al. High frequency of exon 15 deletion in the FANCA gene in Tunisian patients affected with Fanconi anemia disease: implication for diagnosis. Mol Genet Genomic Med. Mar 2014;2(2):160–5. doi:10.1002/mgg3.55

29. Levran O, Erlich T, Magdalena N, et al. Sequence variation in the Fanconi anemia gene FAA. Proc Natl Acad Sci U S A. Nov 25 1997;94(24):13051–6. doi:10.1073/pnas.94.24.13051

30. Faivre L, Guardiola P, Lewis C, et al. Association of complementation group and mutation type with clinical outcome in fanconi anemia. European Fanconi Anemia Research Group. Blood. Dec 15 2000;96(13):4064–70.

31. Fargo JH, Rochowski A, Giri N, Savage SA, Olson SB, Alter BP. Comparison of chromosome breakage in non-mosaic and mosaic patients with Fanconi anemia, relatives, and patients with other inherited bone marrow failure syndromes. Cytogenet Genome Res. 2014;144(1):15–27. doi:10.1159/000366251

32. Ramirez MJ, Pujol R, Trujillo-Quintero JP, et al. Natural gene therapy by reverse mosaicism leads to improved hematology in Fanconi anemia patients. Am J Hematol. Aug 1 2021;96(8):989–999. doi:10.1002/ajh.26234

33. Rickman KA, Lach FP, Abhyankar A, et al. Deficiency of UBE2T, the E2 Ubiquitin Ligase Necessary for FANCD2 and FANCI Ubiquitination, Causes FA-T Subtype of Fanconi Anemia. Cell Rep. Jul 7 2015;12(1):35–41. doi:10.1016/j.celrep.2015.06.014

34. Nicoletti E, Rao G, Bueren JA, et al. Mosaicism in Fanconi anemia: concise review and evaluation of published cases with focus on clinical course of blood count normalization. Ann Hematol. May 2020;99(5):913–924. doi:10.1007/s00277-020-03954-2

35. Dufour C, Pierri F. Modern management of Fanconi anemia. Hematology Am Soc Hematol Educ Program. Dec 9 2022;2022(1):649–657. doi:10.1182/hematology.2022000393

36. Mehta PA, Davies SM, Leemhuis T, et al. Radiation-free, alternative-donor HCT for Fanconi anemia patients: results from a prospective multi-institutional study. Blood. Apr 20 2017;129(16):2308–2315. doi:10.1182/blood-2016-09-743112

37. Murillo-Sanjuan L, Gonzalez-Vicent M, Argiles Aparicio B, et al. Survival and toxicity outcomes of hematopoietic stem cell transplantation for pediatric patients with Fanconi anemia: a unified multicentric national study from the Spanish Working Group for Bone Marrow Transplantation in Children. Bone Marrow Transplant. May 2021;56(5):1213–1216. doi:10.1038/s41409-020-01172-y

38. Peffault de Latour R, Porcher R, Dalle JH, et al. Allogeneic hematopoietic stem cell transplantation in Fanconi anemia: the European Group for Blood and Marrow Transplantation experience. Blood. Dec 19 2013;122(26):4279–86. doi:10.1182/blood-2013-01-479733

39. Ebens CL, DeFor TE, Tryon R, Wagner JE, MacMillan ML. Comparable Outcomes after HLA-Matched Sibling and Alternative Donor Hematopoietic Cell Transplantation for Children with Fanconi Anemia and Severe Aplastic Anemia. Biol Blood Marrow Transplant. Apr 2018;24(4):765–771. doi:10.1016/j.bbmt.2017.11.031

40. Bernard F, Uppungunduri CRS, Meyer S, et al. Excellent overall and chronic graft-versus-host-disease-free event-free survival in Fanconi anaemia patients undergoing matched related- and unrelated-donor bone marrow transplantation using alemtuzumab-Flu-Cy: the UK experience. Br J Haematol. May 2021;193(4):804–813. doi:10.1111/bjh.17418

41. Kahn JM, Brazauskas R, Tecca HR, et al. Subsequent neoplasms and late mortality in children undergoing allogeneic transplantation for nonmalignant diseases. Blood Adv. May 12 2020;4(9):2084–2094. doi:10.1182/bloodadvances.2019000839

42. Rosenberg PS, Socie G, Alter BP, Gluckman E. Risk of head and neck squamous cell cancer and death in patients with Fanconi anemia who did and did not receive transplants. Blood. Jan 1 2005;105(1):67–73. doi:10.1182/blood-2004-04-1652

43. Rio P, Navarro S, Wang W, et al. Successful engraftment of gene-corrected hematopoietic stem cells in non-conditioned patients with Fanconi anemia. Nat Med. Sep 2019;25(9):1396–1401. doi:10.1038/s41591-019-0550-z

44. Lach FP, Singh S, Rickman KA, et al. Esophageal cancer as initial presentation of Fanconi anemia in patients with a hypomorphic FANCA variant. Cold Spring Harb Mol Case Stud. Dec 2020;6(6) doi:10.1101/mcs.a005595

45. Bottega R, Nicchia E, Cappelli E, et al. Hypomorphic FANCA mutations correlate with mild mitochondrial and clinical phenotype in Fanconi anemia. Haematologica. Mar 2018;103(3):417–426. doi:10.3324/haematol.2017.176131

46. Huck K, Hanenberg H, Gudowius S, et al. Delayed diagnosis and complications of Fanconi anaemia at advanced age--a paradigm. Br J Haematol. Apr 2006;133(2):188–97. doi:10.1111/j.1365-2141.2006.05998.x

47. Ramanagoudr-Bhojappa R, Tryon R, Lach FP, et al. FANCA c.3624C>T (p.Ser1208=) is a hypomorphic splice variant associated with delayed onset of Fanconi anemia. Blood Adv. Feb 27 2024;8(4):899–908. doi:10.1182/bloodadvances.2023011888

48. Jaganathan K, Kyriazopoulou Panagiotopoulou S, McRae JF, et al. Predicting Splicing from Primary Sequence with Deep Learning. Cell. Jan 24 2019;176(3):535–548 e24. doi:10.1016/j.cell.2018.12.015

49. Pasquini R, Carreras J, Pasquini MC, et al. HLA-matched sibling hematopoietic stem cell transplantation for fanconi anemia: comparison of irradiation and nonirradiation containing conditioning regimens. Biol Blood Marrow Transplant. Oct 2008;14(10):1141–1147. doi:10.1016/j.bbmt.2008.06.020

50. MacMillan ML, DeFor TE, Young JA, et al. Alternative donor hematopoietic cell transplantation for Fanconi anemia. Blood. Jun 11 2015;125(24):3798–804. doi:10.1182/blood-2015-02-626002

51. Adachi D, Oda T, Yagasaki H, et al. Heterogeneous activation of the Fanconi anemia pathway by patient-derived FANCA mutants. Hum Mol Genet. Dec 1 2002;11(25):3125–34. doi:10.1093/hmg/11.25.3125

52. Castella M, Pujol R, Callen E, et al. Origin, functional role, and clinical impact of Fanconi anemia FANCA mutations. Blood. Apr 7 2011;117(14):3759–69. doi:10.1182/blood-2010-08-299917

53. Garcia-Higuera I, Kuang Y, Denham J, D’Andrea AD. The fanconi anemia proteins FANCA and FANCG stabilize each other and promote the nuclear accumulation of the Fanconi anemia complex. Blood. Nov 1 2000;96(9):3224–30.

