## supplemental material for "Tracking Cytopenias in *FANCA*-deficient Fanconi Anemia"

Short title: Natural History of *FANCA*-deficient Fanconi anemia

Rochelle R. Maxwell, Tamar Berger*, Caroline S. Jiang*, Allana Rosenberg, Ashlyn-Maree Gonzalez, Jodie Odame, Yu-Chien Lin, Francis P. Lach, Jennifer Kennedy, Rebecca Tryon, Frank X. Donovan, Danielle C. Kimble, Shivatheja Soma, Maria I. Cancio, John E. Wagner, Margaret L. MacMillan, Stella M. Davies, Settara C. Chandrasekharappa, Parinda A. Mehta, Farid Boulad, Arleen D. Auerbach and Agata Smogorzewska

**Supplemental files include:**

**Supplemental Figure 1. Examples of individual blood count plots of patients in different groups. A-F.** counts of individuals who had a HSCT before or at 12 years of age. **G-I.** counts of individuals who had a HSCT after 12 years of age. **J.** counts of individuals who had a disease modifying therapy, but not HSCT, **K-N.** counts of individuals who had no disease modifying therapy, and no HSCT

**Supplemental Figure 2. Plots of blood counts in patients with GCSF exposure A.** Platelet count **B.** White Blood Count (WBC) **C.** Hemoglobin **D.** Absolute Neutrophil Count (ANC) **E.** Mean Corpuscular Volume (MCV).

**Supplemental Figure 3. Plots of blood counts while the participants were or were not on androgen. A and F.** Platelet count **B and G.** White Blood Count (WBC) **C and H** Hemoglobin **D and I.** Absolute Neutrophil Count (ANC) **E and J.** Mean Corpuscular Volume (MCV). See also data in Figure 4 where these data are log-transformed.

**Supplemental Table 1: Supplemantal Table 1. Characteristics of the study cohort (n=139) including demographic details, FANCA mutations and their effect, developmental abnormalities, HSCT status, cancer history, vital status and age of last follow up.** UK means unknown. N/A means not applicable *To prevent patient identification, ages are presented by age category in approximately 5-year intervals; statistical analyses were performed using the exact age.

### Supplemental Figure 1A Early HSCT (≤ 12 years)

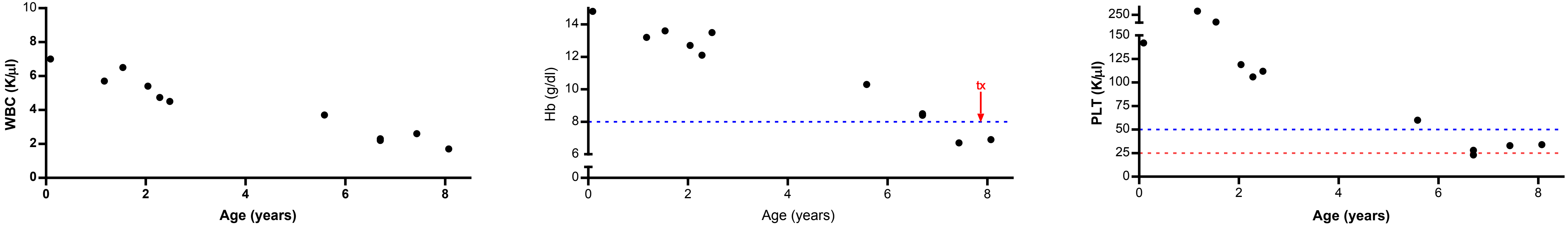


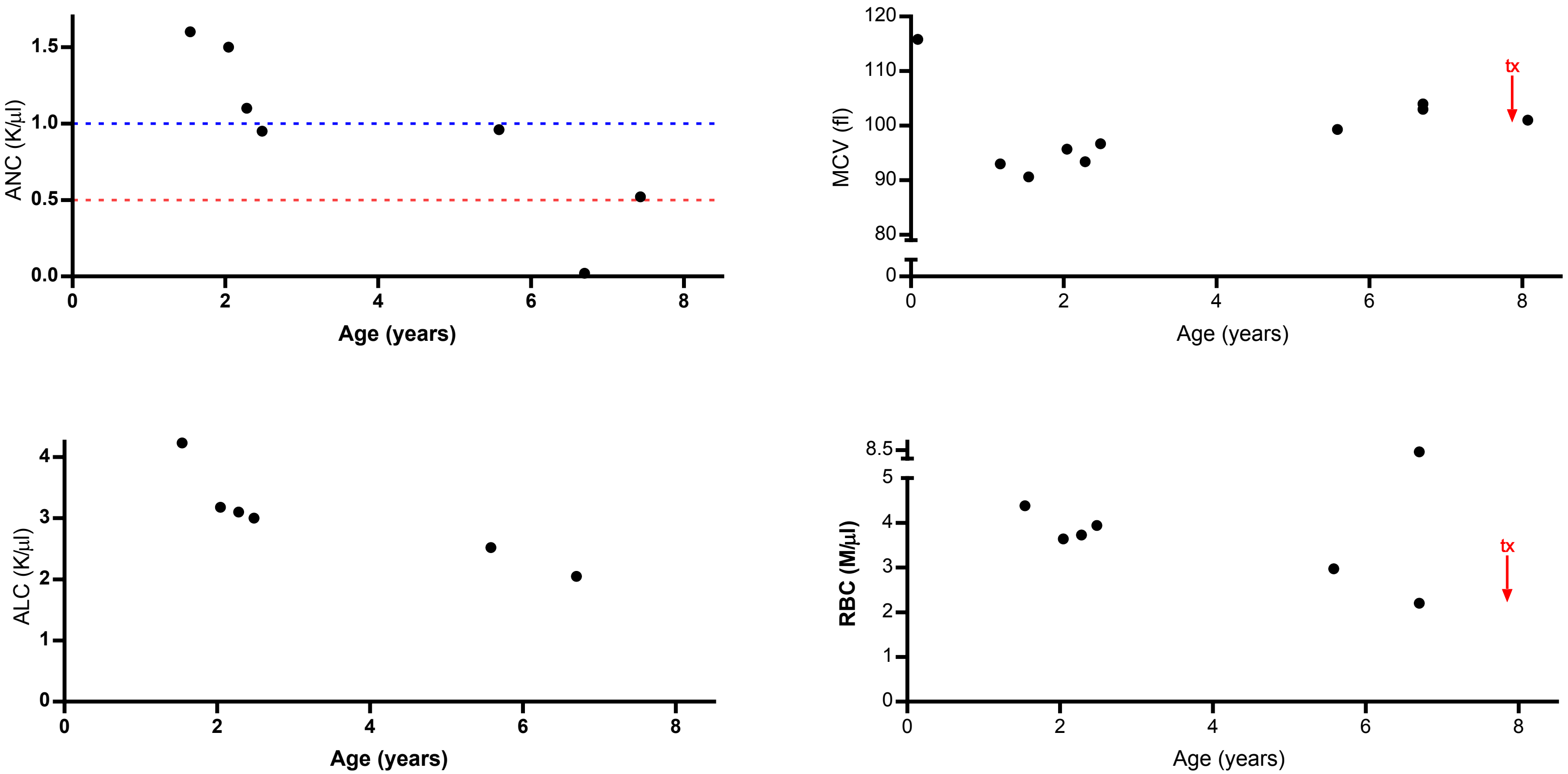


Age at diagnosis: 3 weeks

Congenital anomalies: microcephaly, skeletal, GU, other growth, eye

Age at HSCT: 8.1 years

Mutation 1: In-frame

Mutation 2: Frameshift

| Therapy | Notation |
|---|---|
| Androgen | @ |
| Corticosteroid | # |
| GCSF | % |
| Growth Hormone | ^^ |
| EPO | *e* |
| Transfusion | tx |

### Supplemental Figure 1B Early HSCT (≤ 12 years)

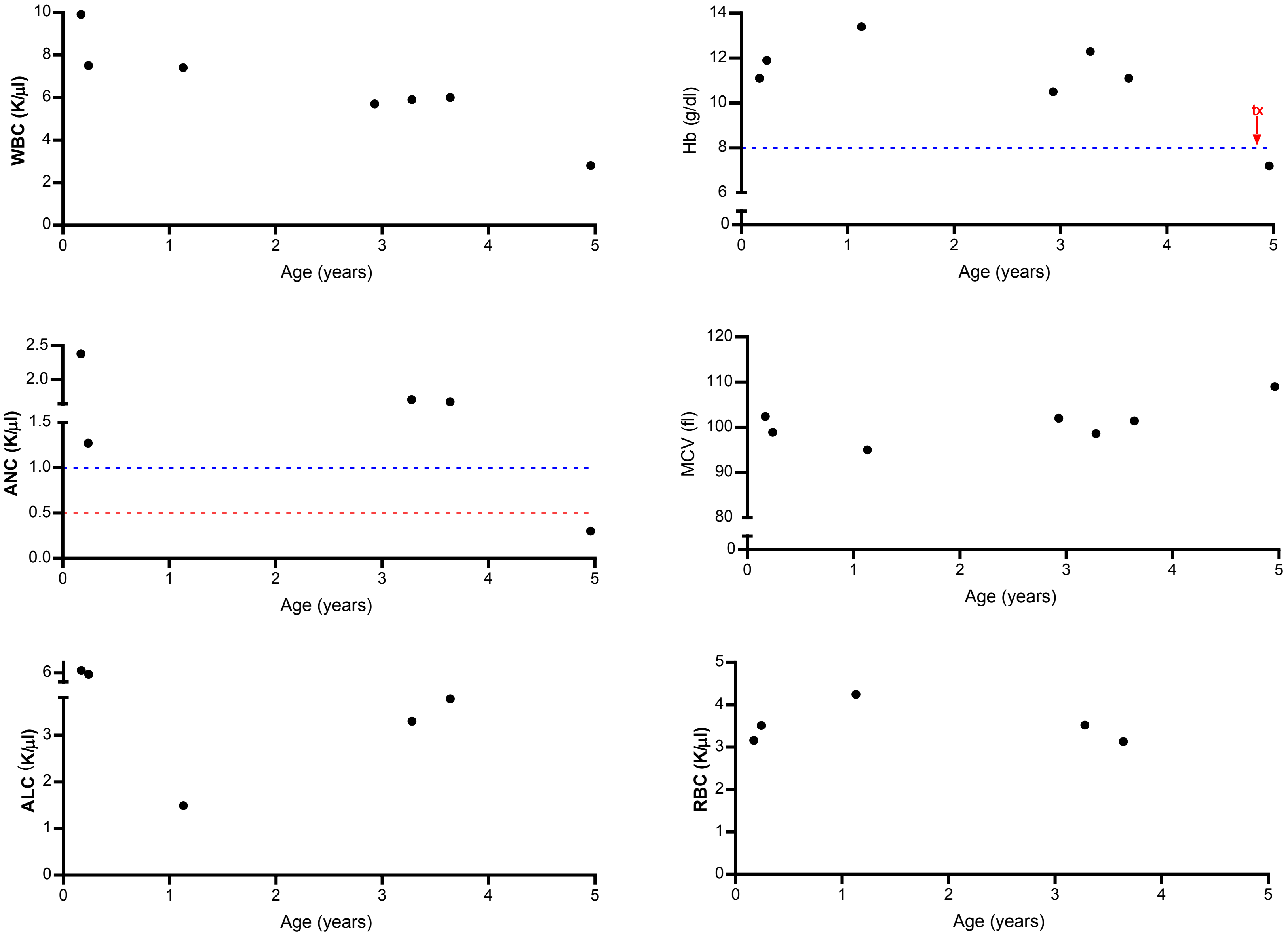


Age at diagnosis: 1 year

Congenital anomalies: skeletal, cardiac, GI, GU, eye, ear

Age at HSCT: 5.2 years

Mutation 1: Frameshift

Mutation 2: Large deletion

### Supplemental Figure 1C Early HSCT (≤ 12 years)

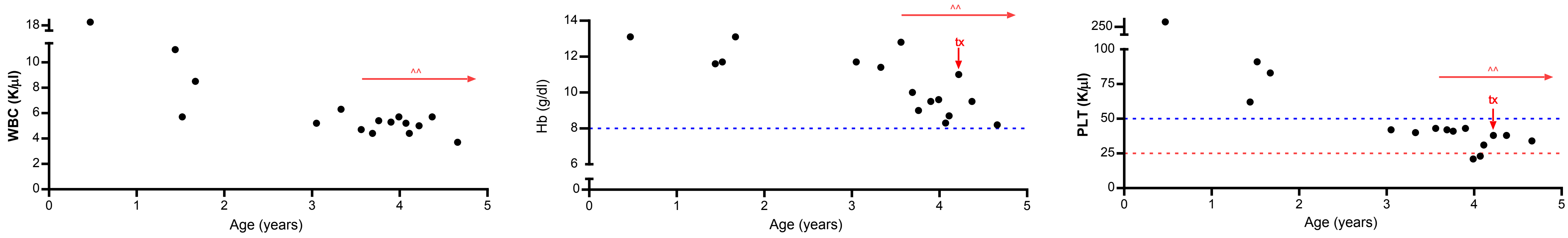


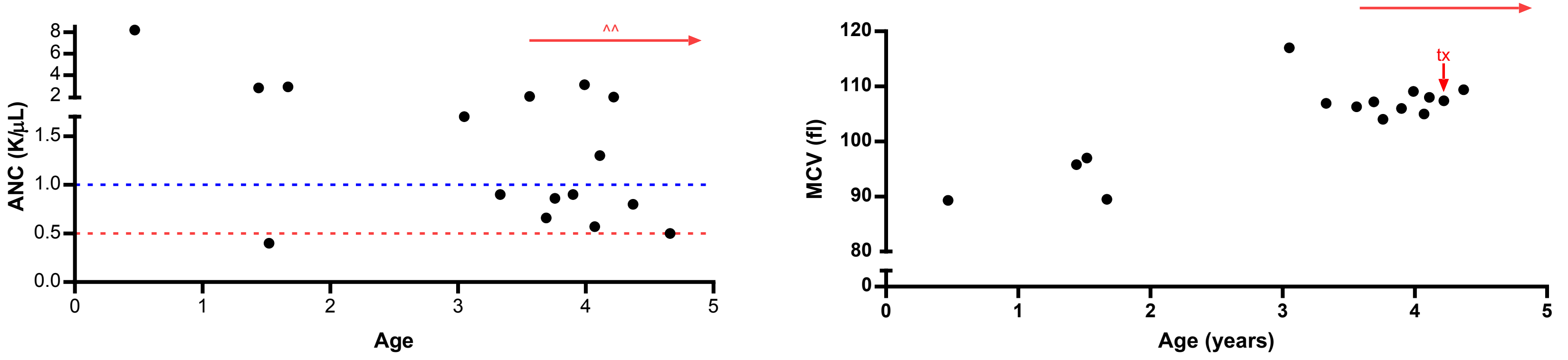


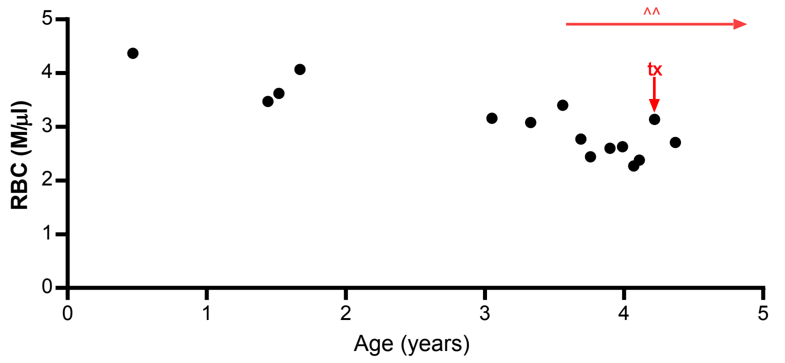


| Therapy | Notation |
|---|---|
| Androgen | @ |
| Corticosteroid | # |
| GCSF | % |
| Growth Hormone | ^^ |
| EPO | *e* |
| Transfusion | tx |

Age at diagnosis: 3 years

Congenital anomalies: skeletal, skin, cardiac, GU, microcephaly, short stature, other growth, eye, ear

Age at HSCT: 4.7 years

Mutation 1: Frameshift

Mutation 2: Frameshift

### Supplemental Figure 1D Early HSCT (≤ 12 years)

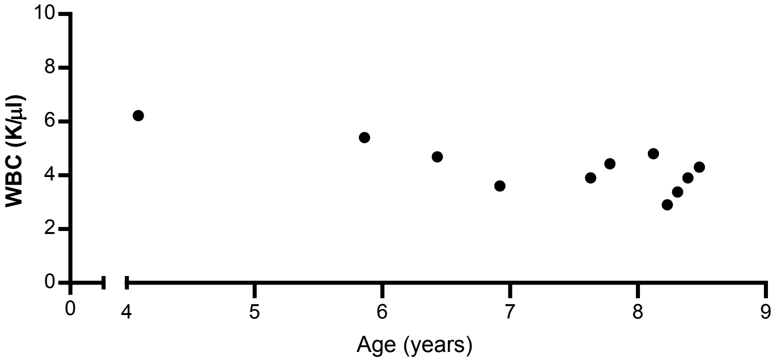


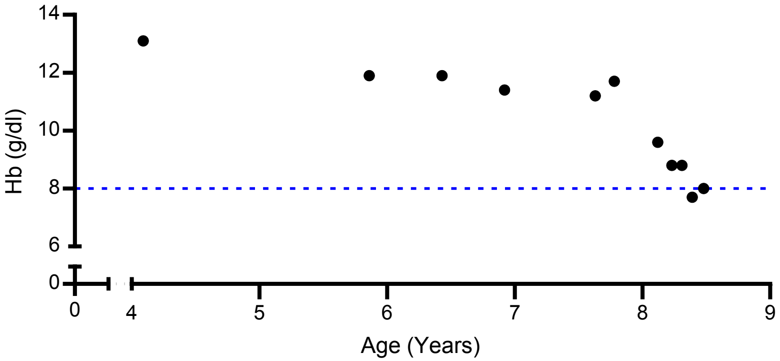


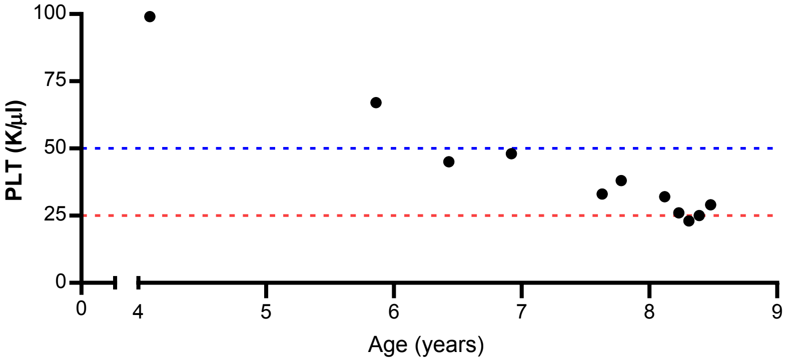


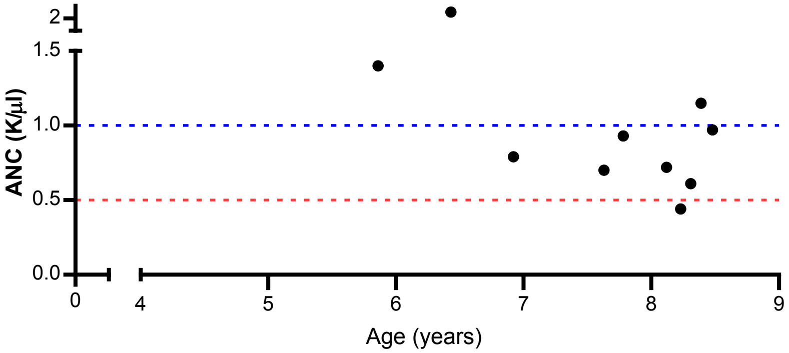


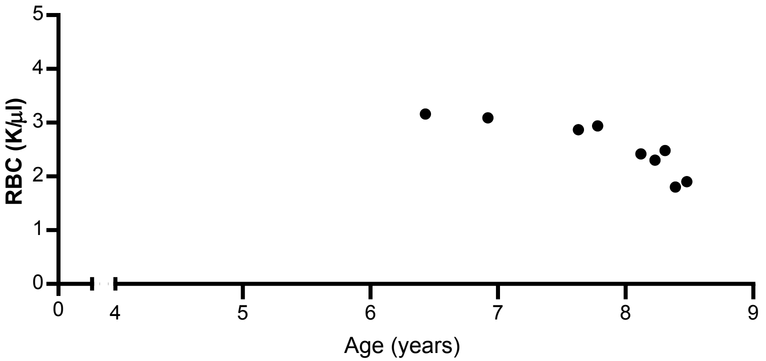


Age at diagnosis: 5 years

Congenital anomalies: skeletal, skin, GI, GU, short stature, other growth

Age at HSCT: 8.7 years

Mutation 1: Nonsense

Mutation 2:  Missense

### Supplemental Figure 1E Early HSCT (≤ 12 years)

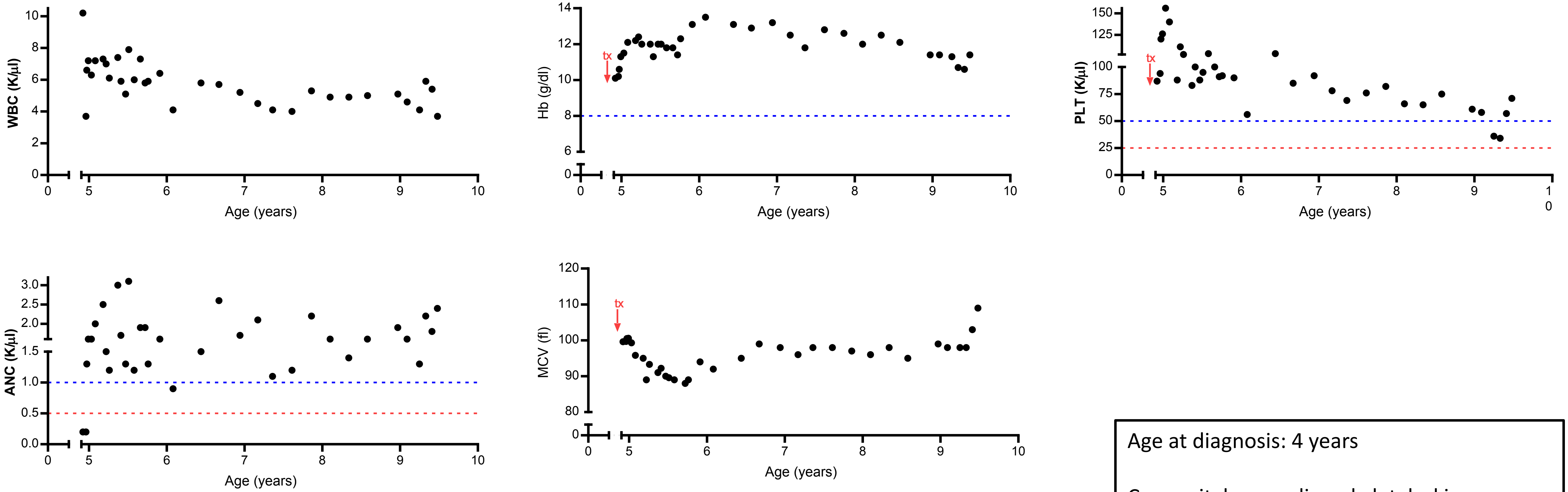


| Therapy | Notation |
| --- | --- |
| Androgen | @ |
| Corticosteroid | # |
| GCSF | % |
| Growth Hormone | ^^ |
| EPO | *e* |
| Transfusion | tx |

Age at diagnosis: 4 years

Congenital anomalies: skeletal, skin, eye

Age at HSCT: 9.5 years

Mutation 1: Splicing

Mutation 2: Splicing

### Supplemental Figure 1F Early HSCT (≤ 12 years)

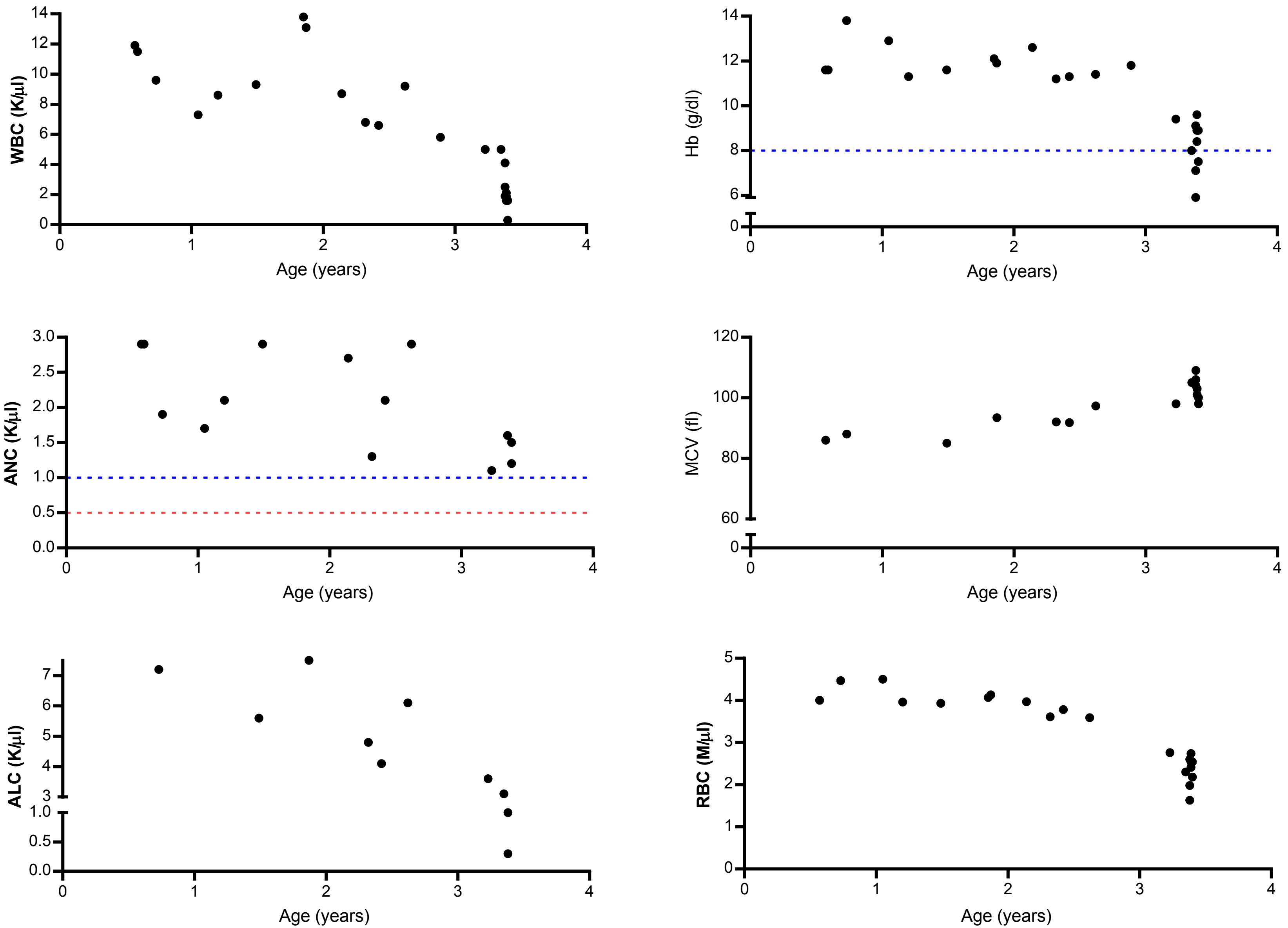


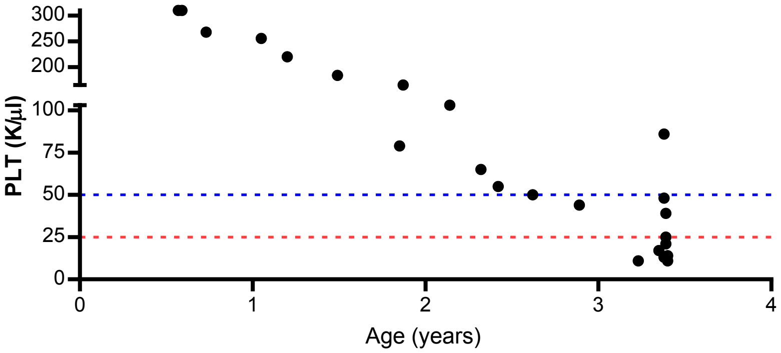


Age at diagnosis: 1 year

Congenital anomalies: microcephaly, other growth

Age at HSCT: 3.4 years

Mutation 1: Frameshift

Mutation 2: Nonsense

### Supplemental Figure 1G Late HSCT (> 12 years)

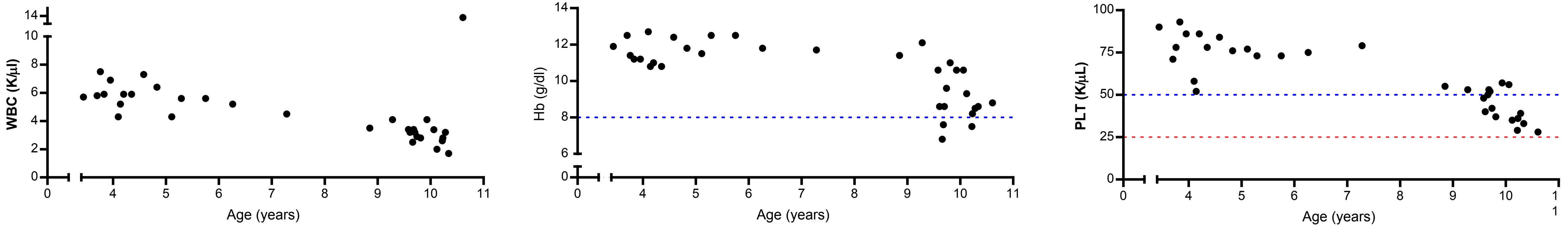


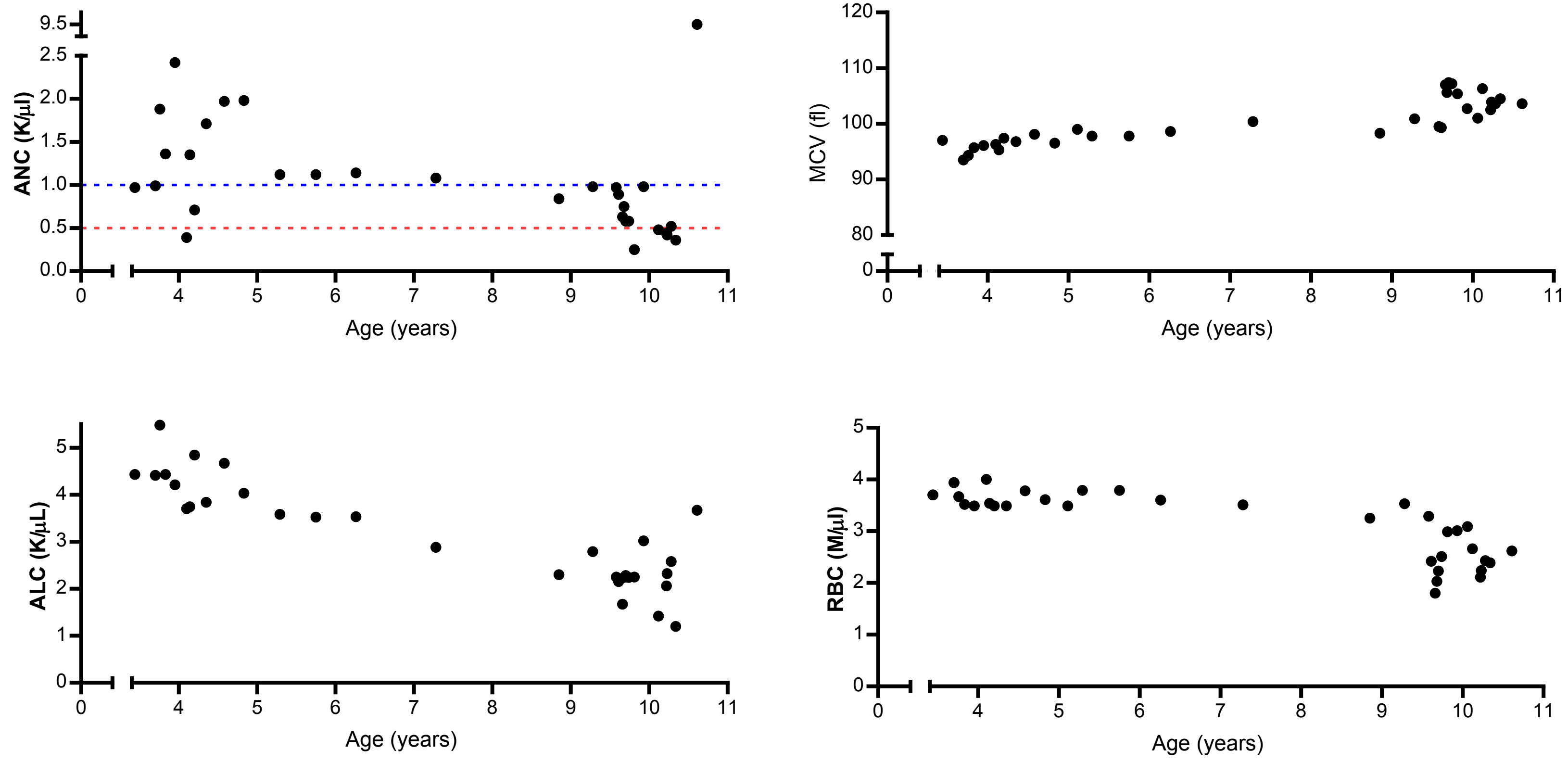


Age at diagnosis: 10 years

Congenital anomalies: skeletal, short stature

Age at HSCT: 13 years

Mutation 1: Missense

Mutation 2: Missense

### Supplemental Figure 1H Late HSCT (> 12 years)

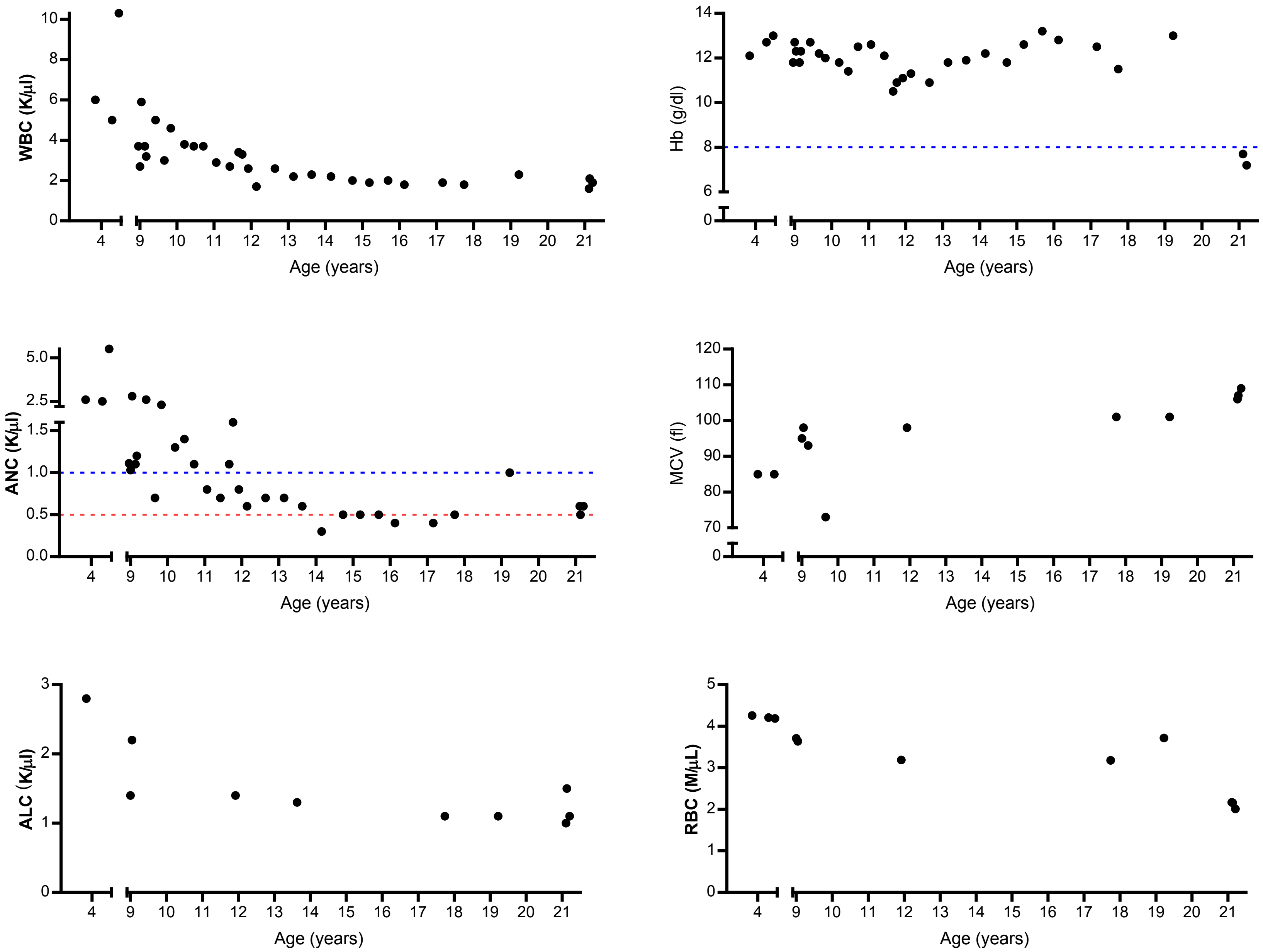


Age at diagnosis: 7 years

Congenital anomalies: skin, GU, short stature

Age at HSCT: 21 years

Mutation 1: Missense

Mutation 2: Splicing

### Supplemental Figure 1I Late HSCT (>12 years)

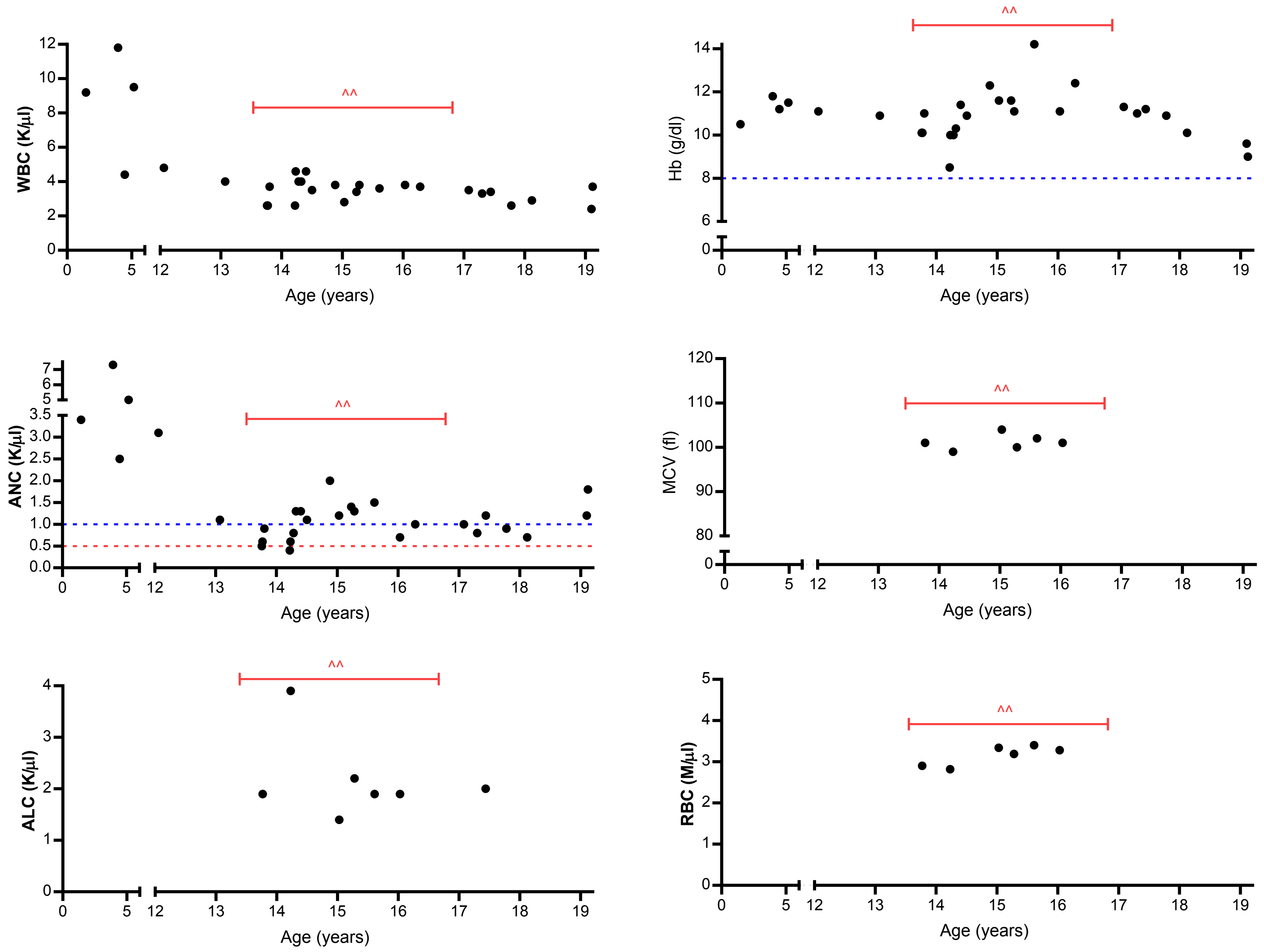


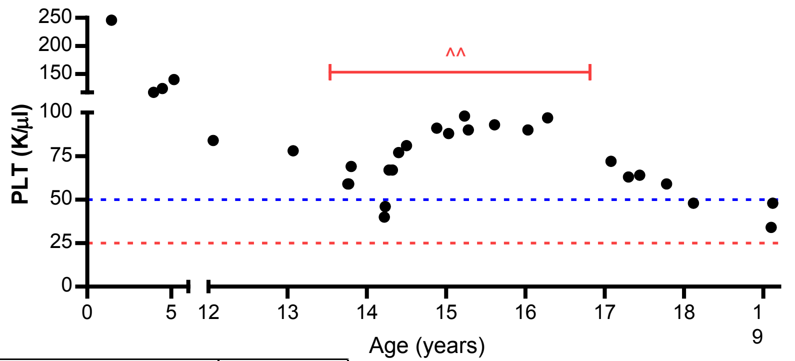


| Therapy | Notation |
|---|---|
| Androgen | @ |
| Corticosteroid | # |
| GCSF | % |
| Growth Hormone | ^^ |
| EPO | *e* |
| Transfusion | tx |

Age at diagnosis: 13 years

Congenital anomalies: skeletal, microcephaly, short stature, eye, ear, hypothyroidism, growth hormone deficiency

Age at HSCT: 21 years

Mutation 1: Nonsense

Mutation 2: Splicing

### Supplemental Figure 1J Disease-modifying therapy without HSCT

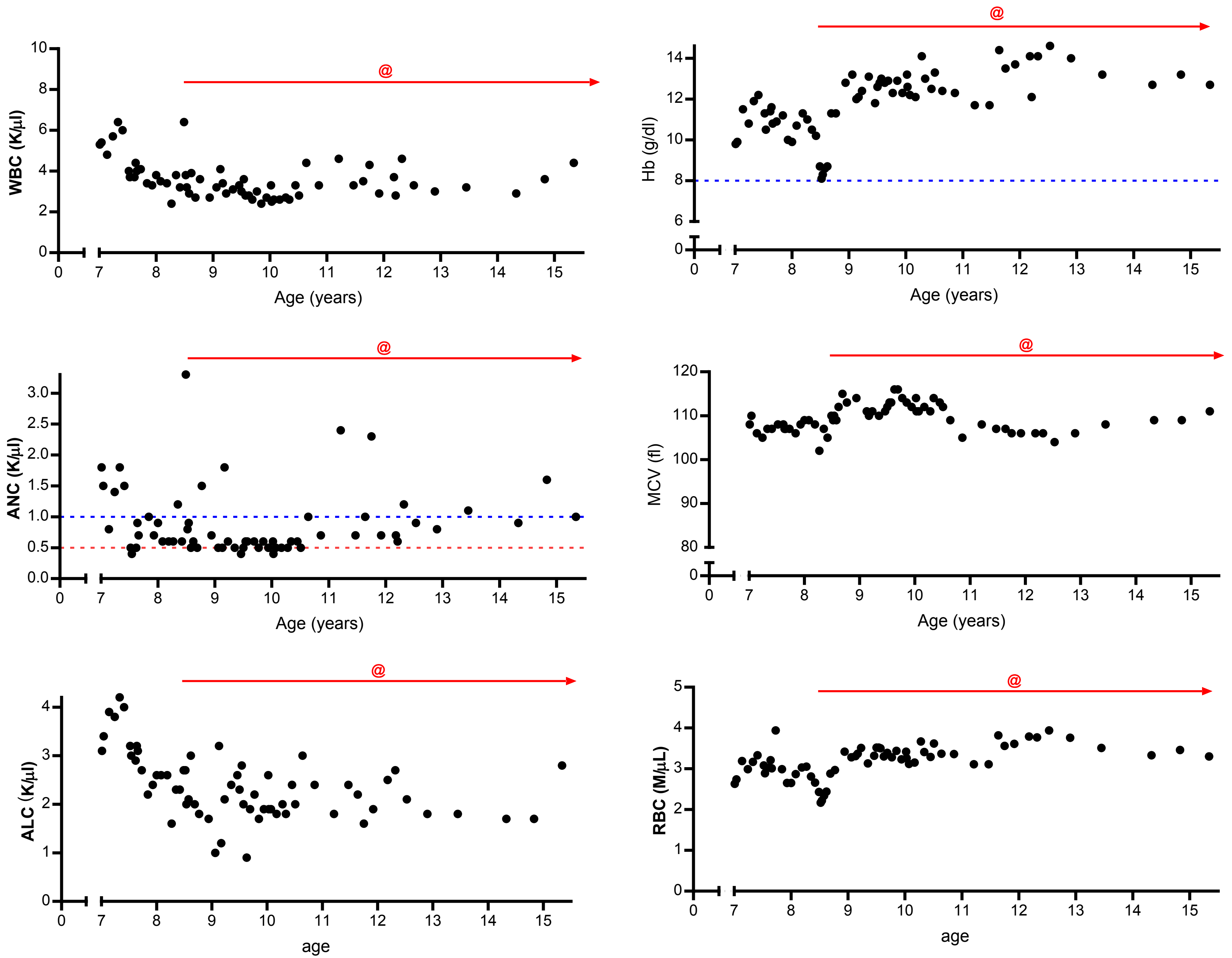


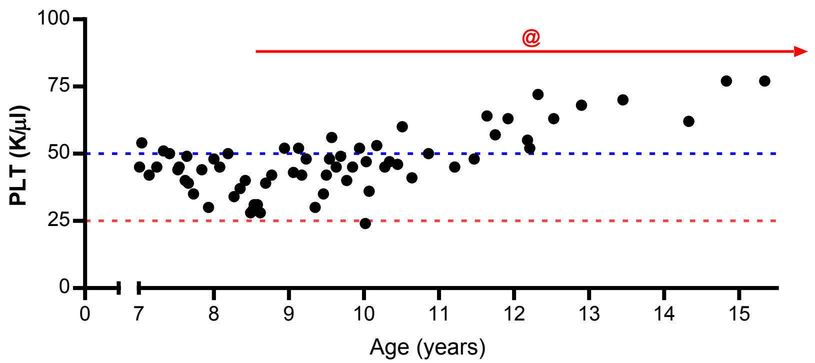


| Therapy | Notation |
|---|---|
| Androgen | **@** |
| Corticosteroid | **#** |
| GCSF | **%** |
| Growth Hormone | **^^** |
| EPO | ***e*** |
| Transfusion | **tx** |

Age at diagnosis: 6 years

Congenital anomalies: skin, microcephaly, short stature, other growth, eye

Age at HSCT: N/A

Mutation 1: Frameshift

Mutation 2: Frameshift

### Supplemental Figure 1K  No disease-modifying therapy without HSCT

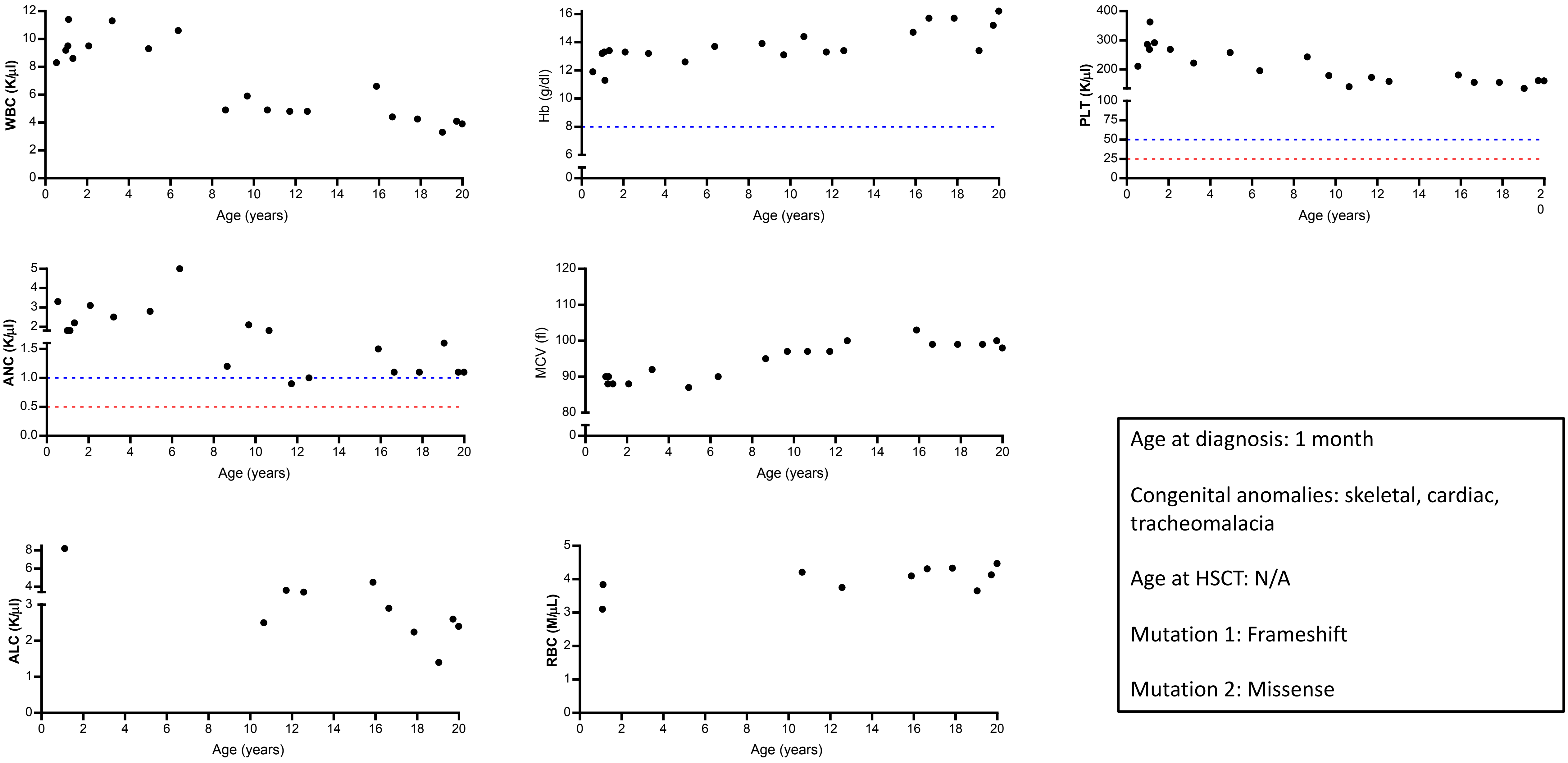

### Supplemental Figure 1L  No disease-modifying therapy without HSCT

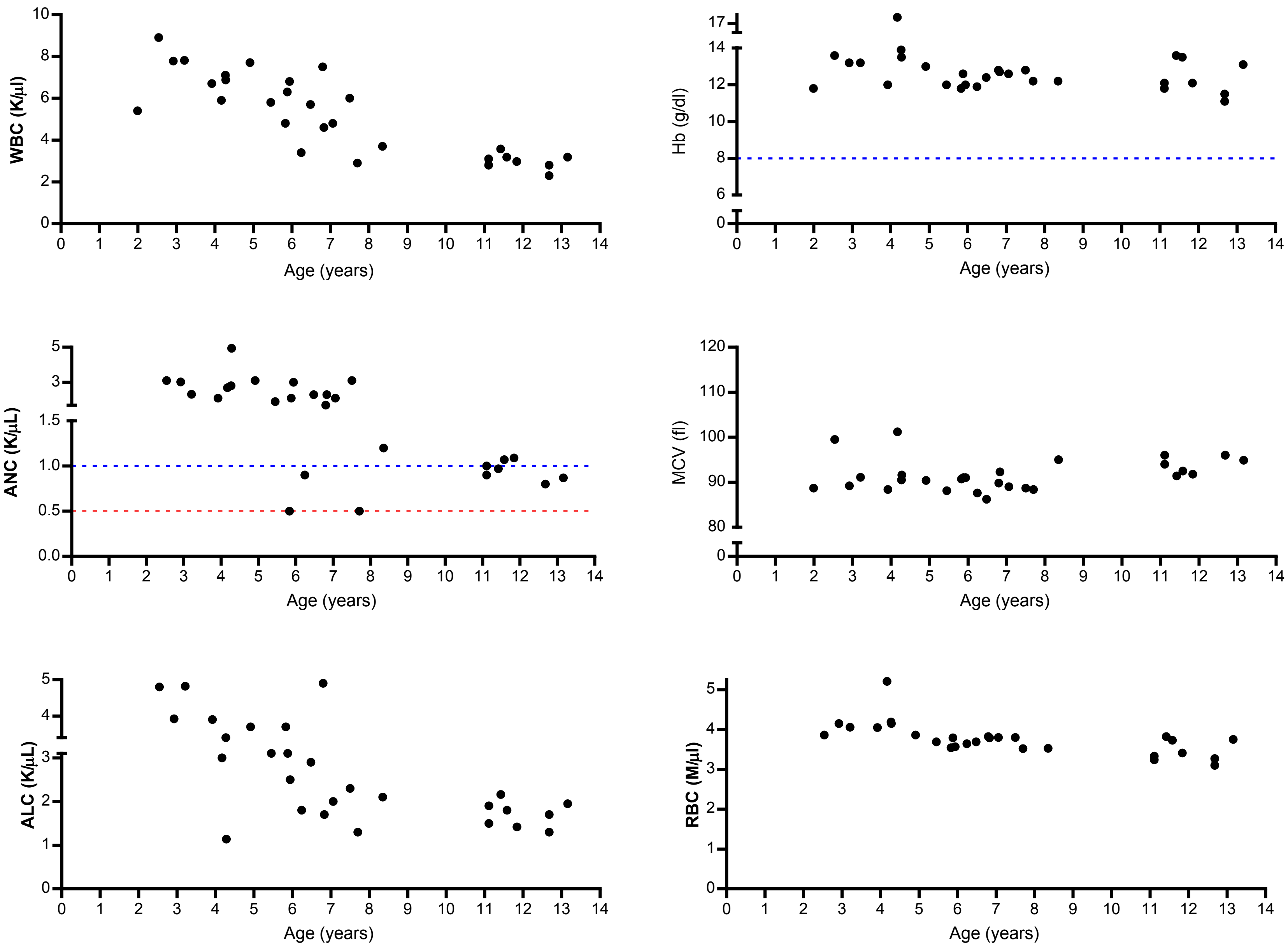


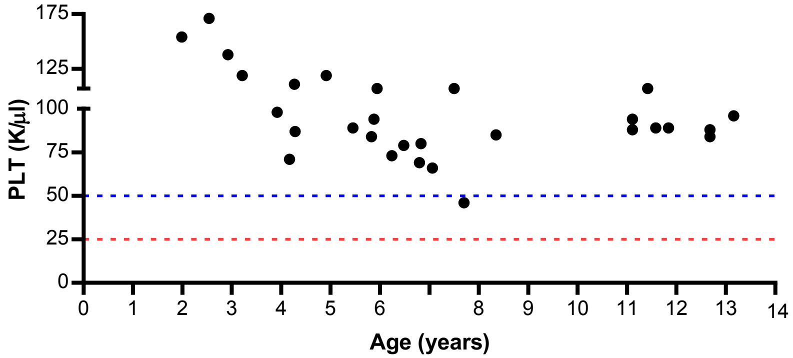


Age at diagnosis: 5 days

Congenital anomalies: skeletal, cardiac, GU, other CNS

Age at HSCT: N/A

Mutation 1: Missense

Mutation 2: Large deletion

### Supplemental Figure 1M  No disease-modifying therapy without HSCT

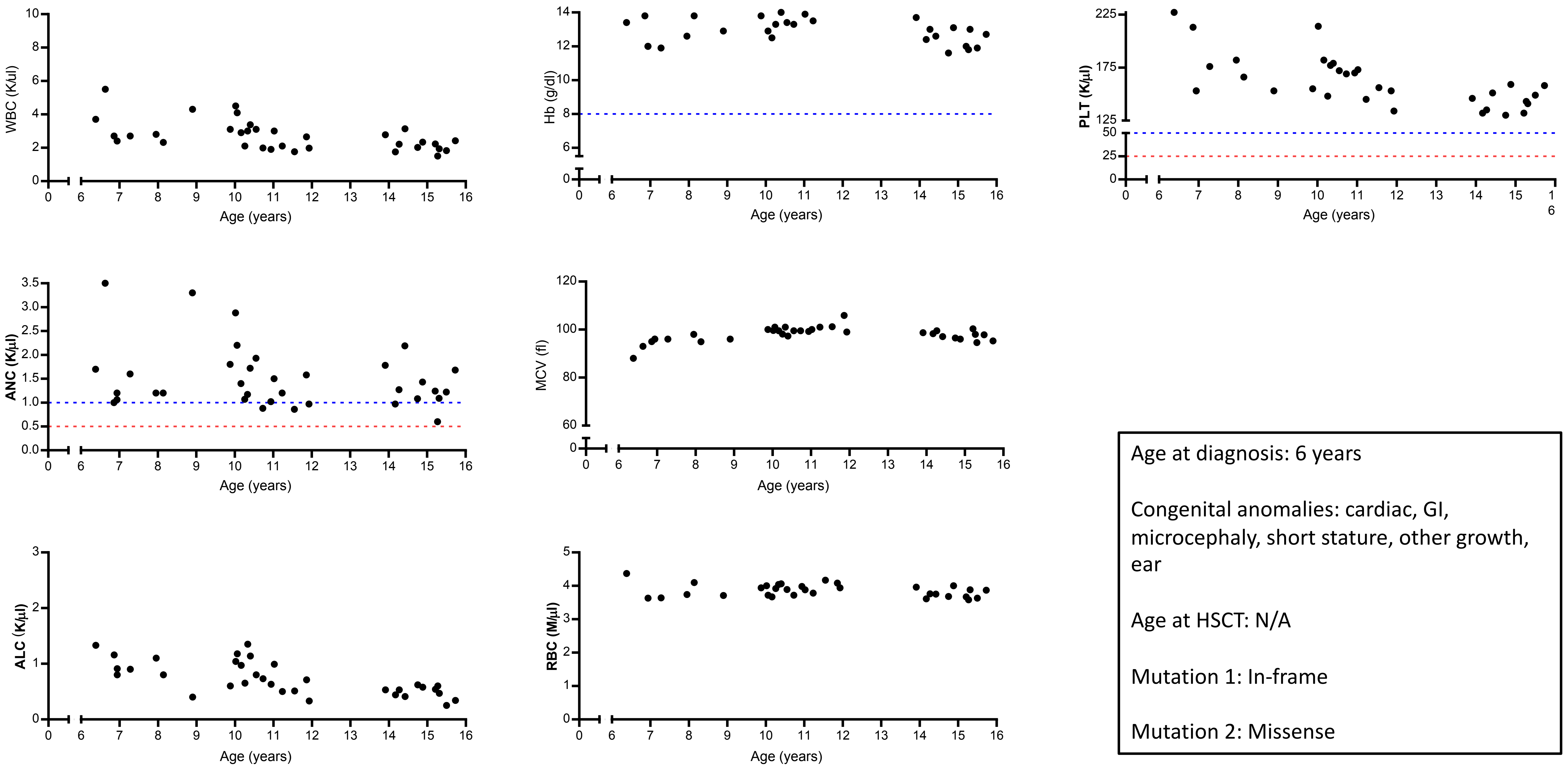


Age at diagnosis: 6 years

Congenital anomalies: cardiac, GI, microcephaly, short stature, other growth, ear

Age at HSCT: N/A

Mutation 1: In-frame

Mutation 2: Missense

### Supplemental Figure 1N  No disease-modifying therapy without HSCT

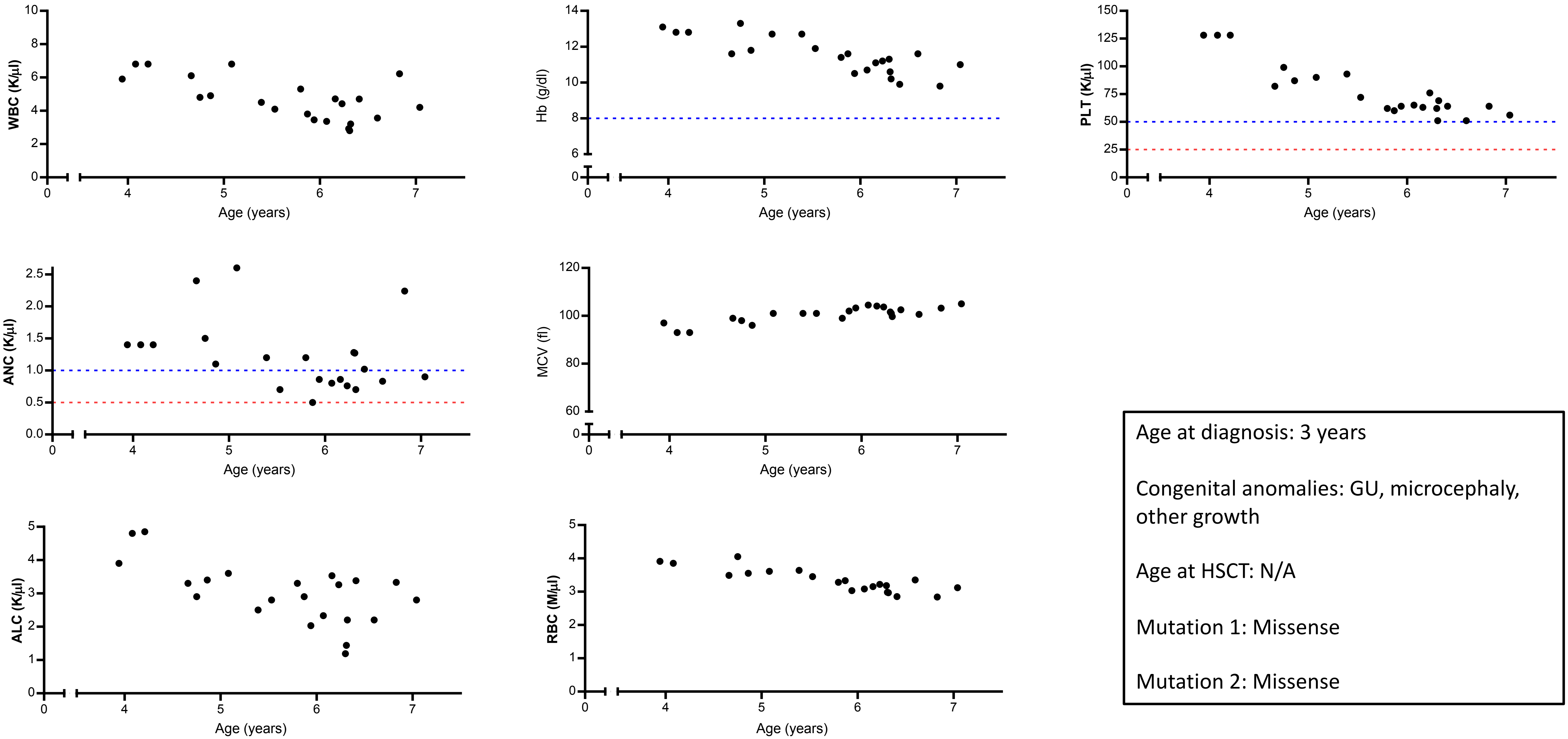


Age at diagnosis: 3 years

Congenital anomalies: GU, microcephaly, other growth

Age at HSCT: N/A

Mutation 1: Missense

Mutation 2: Missense

Maxwell et al. Figure S2

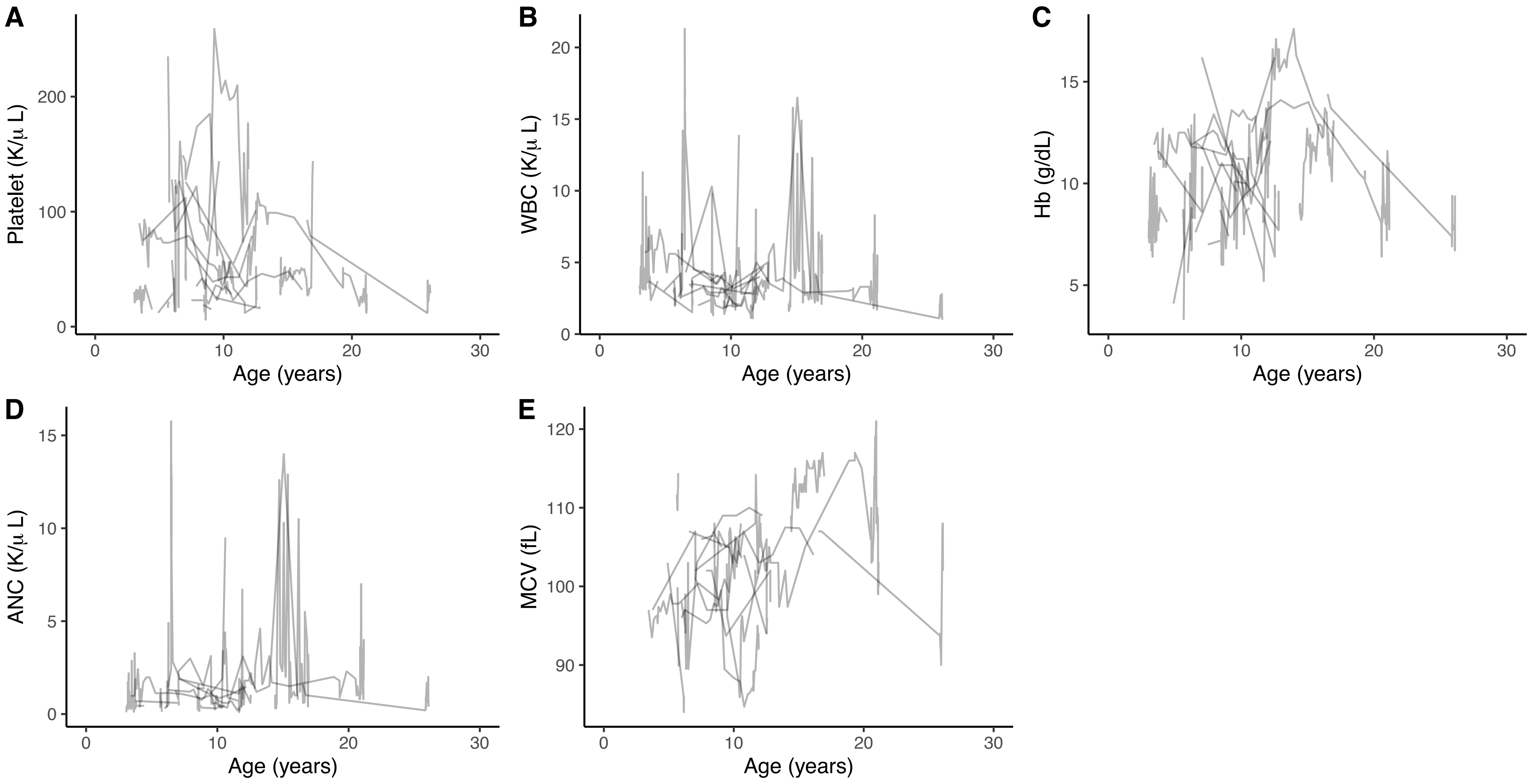

Maxwell et al. Figure S3

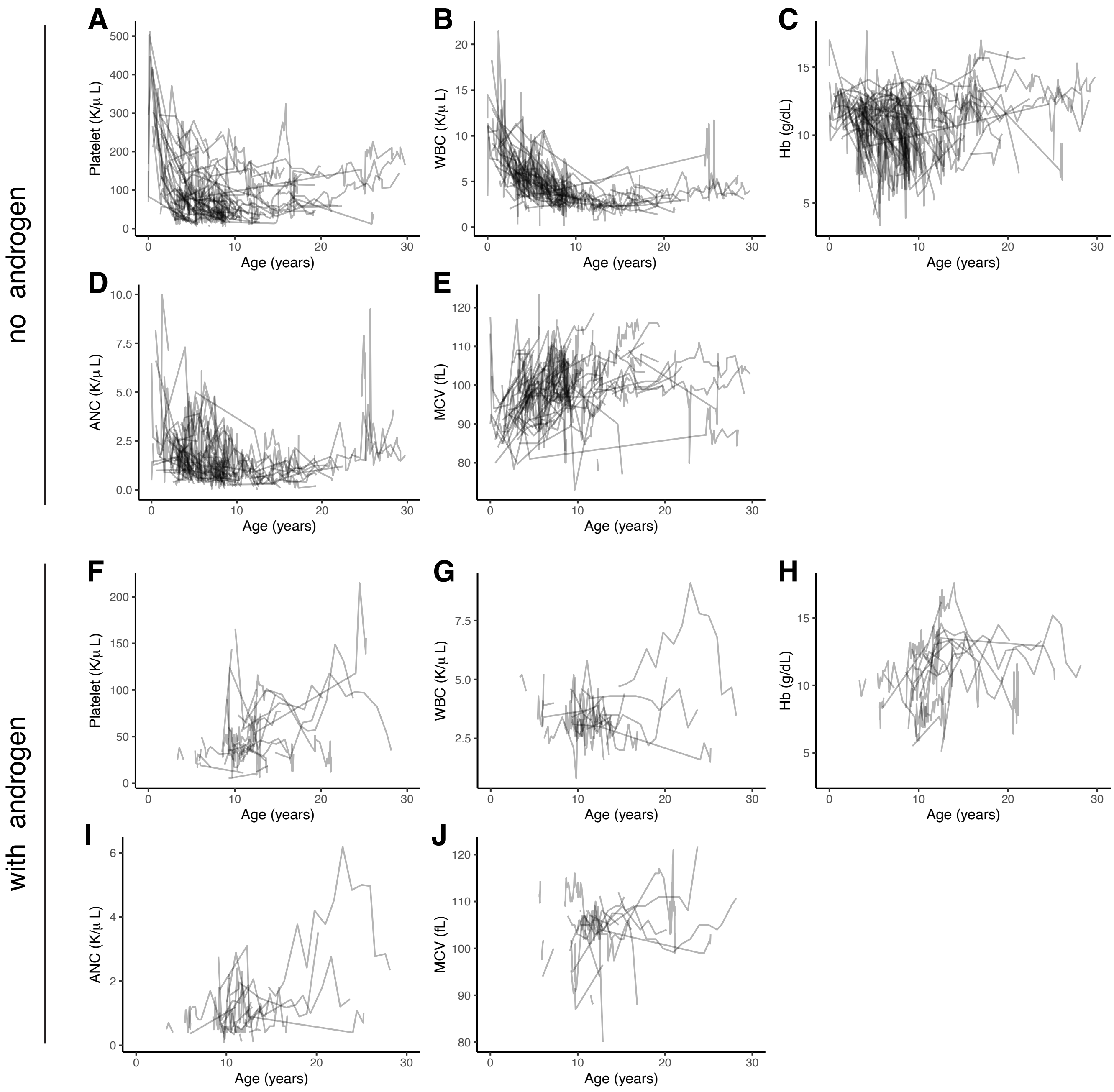

Supplemantal Table 1. Characteristics of the study cohort (n=139) including demographic details, FANCA mutations and their effect, developmental abnormalities, HSCT status, cancer history, vital status and age of last follow up. UK means unknown. N/A means not applicable *To prevent patient identification, ages are presented by age category in approximately 5-year intervals; statistical analyses were performed using the exact age. .

| | Age at diagnosis | Race | Ethnicity | Gender | Mutation 1 | | | | | | Mutation 2 | | | | | | Congenital Abnormalities | Allogenic HSCT | MDS/AML/solid tumors | | Vital Status | Age at Death | Cause of death | Age at last follow up |
|---|---|---|---|---|---|---|---|---|---|---|---|---|---|---|---|---|---|---|---|---|---|---|---|---|
| | years, unless otherwise specified | | Latino: Yes/No/UK | F /M | Type | Effect | Effect_coded | Exon | Variant (NM_000135.2, hg19) | Protein | Type | Effect | Effect_coded | Exon | Variant (NM_000135.2, hg19 | Protein | | Age of first transplant (years)/ None/ UK | Age of diagnosis (years)/ None/ UK | Type (N/A if no cancer) | Alive/ Deceased/ UK | years | | years |
| 1 | 5-9.99 | White | UK | F | Substitution | Splicing | 4 | i6 | c.597-1G>C | r.spl | Substitution | Splicing | 4 | i6 | c.597-1G>C | r.spl | skin, cardiac | UK | None | N/A | Deceased | 5-9.99 | UK | |
| 2 | 5-9.99 | White | No | M | Substitution | Splicing | 4 | i20 | c.1827-1G>A | r.spl | Deletion | Large Deletion | 5 | 1_5 | g.89876498_89909748del33251 | Genomic deletion | skeletal, eye, ear | UK | None | N/A | Deceased | 5-9.99 | Other (Multisystem failure) | |
| 3 | <1 | White | No | M | Deletion | In-frame | 3 | 38 | c.3788_3790delTCT | p.F1263del | Deletion | Frameshift | 2 | 41 | c.4069_4082del14 | p.A4071Lfs*63 | skeletal, cardiac, GU, short stature | None | None | N/A | Deceased | 1-4.99 | Other (sepsis) | |
| 4 | 1-4.99 | White | No | M | Substitution | Nonsense | 1 | 29 | c.2840C>G | p.S947* | Deletion | Large Deletion | 5 | 15_21 | g.89841018_89856681del15664 | Genomic deletion | skeletal, skin, GU, short stature | 1-4.99 | 20-24.99 | Thyroid carcinoma | Alive | | | 25-29.99 |
| 5 | 1-4.99 | White | No | M | Substitution | Nonsense | 1 | 18 | c.1645C>T | p.Q549* | Duplication | Frameshift | 2 | 29 | c.2812_2830dup | p.D944Gfs*5 | skin, short stature | 1-4.99 | 1-4.99 | AML | UK | | | 15-19.99 |
| 6 | 5-9.99 | White | Yes | M | Deletion | Splicing | 4 | i42 | c.4261-19_4261-12del | r.spl | Deletion | Splicing | 4 | i42 | c.4261-19_4261-12del | r.spl | skeletal, GU | 10-14.99 | None | N/A | Alive | | | 30-34.99 |
| 7 | 10-14.99 | White | No | F | UK | Undetermined | 6 | | | | UK | Undetermined | 6 | | | | skeletal | None | Cancers starting at 30-34.99 years of age | Multiple skin cancers (SCC, basal; 34 yo), hypopharyngeal SCC, 34 yo), breast (38 yo) | Alive | | | 35-39.99 |
| 8 | 10-14.99 | White | No | F | Deletion | In-frame | 3 | 38 | c.3788_3790delTCT | p.F1263del | UK | Undetermined | 6 | | | | skeletal, GI, other growth | None | None | N/A | Alive | | | 40-44.99 |
| 9 | 1-4.99 | Asian | No | M | Deletion | Large Deletion | 5 | 3_43 | g.88257445_88409236del151792 | Genomic deletion | Substitution | Splicing | 4 | i20 | c.1777-1G>C | r.spl | skeletal, eye, ear | 5.67 | None | N/A | UK | | | 15-19.99 |
| 10 | 1-4.99 | White | Yes | M | UK | Undetermined | 6 | | | | UK | Undetermined | 6 | | | | skeletal, skin, microcephaly, eye, ear | 1-4.99 | None | N/A | Alive | | | 25-29.99 |
| 11 | 1-4.99 | White | No | M | Deletion | Large Deletion | 5 | 22_30 | c.1901-?_2981+?del | Genomic deletion | Substitution | Missense | 0 | 34 | c.3391A>G | p.T1131A | skeletal, skin, GU, short stature, other growth | None | 25-29.99 | Hepatocellular carcinoma | Alive | | | 25-29.99 |
| 12 | 1-4.99 | White | No | F | Substitution | Missense | 0 | 35 | c.3490C>T | p.P1164S | Substitution | Nonsense (start-loss) | 1 | 1 | c.2T>C | p.M1? | skeletal, skin, GU, microcephaly, short stature, ear | 5-9.99 | UK | UK | Alive | | | 25-29.99 |
| 13 | 1-4.99 | White | No | F | Substitution | Nonsense (stop-gain) | 1 | 2 | c.154C>T | p.R52* | Substitution | Missense | 0 | 27 | c.2534T>C | p.L845P | skeletal, skin, short stature | 5.51 | None | N/A | Alive | | | 25-29.99 |
| 14 | 5-9.99 | White | No | F | Substitution | Nonsense (stop-gain) | 1 | 27 | c.2529C>A | p.Y843* | Substitution | Splicing | 4 | i31 | c.3066+1G>A | r.spl | skin | 10-14.99 | UK | UK | UK | | | 15-19.99 |
| 15 | 5-9.99 | White | No | F | Deletion | Large Deletion | 5 | 27_43 | g.89757989_89834621del76633 | Genomic deletion | Substitution | Missense | 0 | 35 | c.3490C>T | p.P1164S | skeletal, ear | 15-19.99 | 25-29.99 | SCC | Deceased | 30-34.99 | Cancer | |
| 16 | 5-9.99 | Other | Yes | M | Duplication | Frameshift | 2 | 25 | c.2233dupT | p.W745Lfs*49 | Deletion | Large Deletion | 5 | 16_43 | g.89800194_89850461del50268 | Genomic deletion | skeletal, GU, microcephaly, other | 5-9.99 | None | N/A | Deceased | 10-14.99 | Post BMT complications | |
| 17 | 10-14.99 | White | No | F | Substitution | Splicing | 4 | i20 | c.1827-1G>A | r.spl | Deletion | Frameshift | 2 | 39 | c.3920delA | p.Q1307Rfs*2 | skeletal, skin | UK | UK | UK | Alive | | | 35-39.99 |

| | | | | | | | | | | | | | | | | | | | | | | | | |
|---|---|---|---|---|---|---|---|---|---|---|---|---|---|---|---|---|---|---|---|---|---|---|---|---|
| 18 | 5-9.99 | White | Yes | F | Deletion | Frameshift | 2 | 13 | c.1115_1118delTTGG | p.V372Afs*42 | Substitution | Nonsense (Stop-gain) | 1 | 1 | c.65G>A | p.W22* | none | 25-29.99 | None | N/A | Deceased | 25-29.99 | Post BMT complications (viral infection) | |
| 19 | <1 | White | No | F | Deletion | Frameshift | 2 | 12 | c.1034_1035delAG | p.E345Vfs*63 | Deletion | Large Deletion | 5 | 28_32 | c.2602-?_3239+?del | Genomic deletion | skeletal, GI, eye | 5-9.99 | UK | UK | Alive | | | 20-24.99 |
| 20 | <1 | White | No | M | Deletion | In-frame | 3 | 38 | c.3788_3790delTCT | p.F1263del | Duplication | Frameshift | 2 | 38 | c.3813dupA | p.H1272Tfs*6 | skeletal, GU, microcephaly, other growth, eye | 5-9.99 | None | N/A | Alive | | | 20-24.99 |
| 21 | 5-9.99 | White | No | M | Substitution | Nonsense (stop-gain) | 1 | 23 | c.2021C>A | p.S674* | Deletion | Large Deletion | 5 | 30_43 | g.89801519_89827804del26286 | Genomic deletion | skeletal, GU | 5-9.99 | UK | UK | UK | | | 15-19.99 |
| 22 | <1 | White | No | M | Substitution | Nonsense (stop-gain) | 1 | 1 | c.65G>A | p.W22* | Deletion | Large Deletion | 5 | 1_6 | g.89871248_89898248del27001 | Genomic deletion | skeletal, GU, microcephaly | 5-9.99 | UK | UK | Alive | | | 20-24.99 |
| 23 | <1 | White | No | M | Duplication | Frameshift | 2 | 36 | c.3558dupG | p.R1187Efs*28 | Substitution | Missense | 0 | 25 | c.2290C>T | p.R764W | skeletal, cardiac, tracheomalacia | None | None | N/A | Alive | | | 20-24.99 |
| 24 | 5-9.99 | White | No | F | Deletion | Large Deletion | 5 | 39_43 | c.3829-?4368+?del | Genomic deletion | Substitution | Nonsense (stop-gain) | 1 | 9 | c.811C>T | p.Q271* | skin, GU, microcephaly, eye | 5-9.99 | UK | MDS | Alive | | | 25-29.99 |
| 25 | 10-14.99 | White | UK | F | Deletion | Large Deletion | 5 | 31_43 | g.89803856_89824374del20519 | Genomic deletion | Substitution | Splicing | 4 | 36 | c.3624C>T | p.S1208S results in r.spl | skin | 20-24.99 | None | N/A | Alive | | | 30-34.99 |
| 26 | <1 | White | No | M | Substitution | Missense | 0 | 29 | c.2852G>A | p.R951Q | Substitution | Splicing | 4 | i39 | c.3934+2T>C | r.spl | skeletal, GU, microcephaly | 10-14.99 | None | N/A | Alive | | | 20-24.99 |
| 27 | 10-14.99 | White | No | M | Substitution | Undetermined | 6 | 31 | c.2989A>T | p.S997C | Substitution | Missense | 0 | 31 | c.3043G>A | p.E1015K | skeletal, GU, eye | 15-19.99 | None | N/A | Deceased | 15-19.99 | Post BMT complications (septic shock) | |
| 28 | 5-9.99 | Other | Yes | M | Substitution | Nonsense (start-loss) | 1 | 1 | c.1A>C | p.M1? | Substitution | Nonsense (start-loss) | 1 | 1 | c.2T>C | p.M1? | GU | 5-9.99 | 5-9.99 | MDS | Deceased | 5-9.99 | Post BMT complications | |
| 29 | <1 | White | No | F | Deletion | Large Deletion | 5 | 18_24 | g.89830232 - 89837758 | Genomic deletion | Substitution | Missense | 0 | 34 | c.3391A>G | p.T1131A | skeletal, cardiac, ear | 10-14.99 | UK | UK | Alive | | | 20-24.99 |
| 30 | 1-4.99 | Other | Yes | M | Deletion | Large Deletion | 5 | 1_43 | c.-42-?_4368+?del | Genomic deletion | Deletion | In-frame | 3 | 38 | c.3788_3790delTCT | p.F1263del | skeletal, skin, GU, eye | 5-9.99 | 15-19.99 | Oral SCC | Deceased | 15-19.99 | Cancer | |
| 31 | 10-14.99 | White | No | F | Substitution | Missense | 0 | 34 | c.3391A>G | p.T1131A | Substitution | Missense | 0 | 28 | c.2606A>C | p.Q869P | skeletal, short stature | 10-14.99 | None | N/A | Deceased | 10-14.99 | Post BMT complications | |
| 32 | 5-9.99 | White | No | F | Deletion | Large Deletion | 5 | 1_43 | c.-42-?_4368+?del | Genomic deletion | Substitution | Missense | 0 | 29 | c.2852G>A | p.R951E | skeletal, skin, GI, GU, other growth, ear | 5-9.99 | 20-24.99 | Vulvar cancer | Alive | | | 25-29.99 |
| 33 | 1-4.99 | White | No | M | Deletion | Large Deletion | 5 | 1_43 | c.-42-?_4368+?del | Genomic deletion | Substitution | Missense | 0 | 29 | c.2852G>A | p.R951E | skeletal, GU, ear | 5-9.99 | None | N/A | Alive | | | 20-24.99 |
| 34 | 10-14.99 | Black/ African American | No | F | Substitution | Missense | 0 | 33 | c.3339C>G | p.N1113K | Substitution | Nonsense (start-loss) | 1 | 1 | c.1A>T | p.M1? | skeletal, skin, GU | 10-14.99 | None | N/A | Deceased | 10-14.99 | Post BMT complications | |
| 35 | 1-4.99 | White | No | M | Deletion | Frameshift | 2 | 13 | c.1115_1118delTTGG | p.V372Afs*42 | Duplication | Frameshift | 2 | 36 | c.3558dupG | p.R1187Efs*28 | skeletal, cardiac, GI, GU, other growth, eye | 1-4.99 | None | N/A | Alive | | | 20-24.99 |
| 36 | 5-9.99 | White | No | F | Deletion | Undetermined | 6 | 39 | c.3846_3856del11 | p.K1283Rfs*26 | Substitution | Nonsense (stop-gain) | 1 | 32 | c.3130C>T | p.Q1044* | skeletal, skin, GI, GU, short [illegible] | 5-9.99 | UK | UK | Alive | | | 25-29.99 |
| 37 | 5-9.99 | White | No | M | Deletion | In-frame | 3 | 38 | c.3788_3790delTCT | p.F1263del | Substitution | Splicing | 4 | i22 | c.2015-1G>T | r.spl | skeletal, skin, GI, GU, other growth, ear | 5-9.99 | 5-9.99; 25-29.99 | AML / MDS (8); oral cavity cancer (26) | Alive | | | 25-29.99 |

| | | | | | | | | | | | | | | | | | | | | | | | | |
|---|---|---|---|---|---|---|---|---|---|---|---|---|---|---|---|---|---|---|---|---|---|---|---|---|
| 38 | 5-9.99 | White | No | F | Deletion | Frameshift | 2 | 37 | c.3760_3761delGA | p.E1255Rfs*12 | Substitution | Missense | 0 | 29 | c.2831A>C | p.D944A | skeletal, skin, microcephaly, other growth, eye, ear | 5-9.99 | 5-9.99; 10-14.99 | MDS (6); AML (10.3) | Alive | | | 25-29.99 |
| 39 | 1-4.99 | White | No | M | Deletion | In-frame | 3 | 36 | c.3520_3522delTGG | p.W1174del | Substitution | Splicing | 4 | i20 | c.1827-1G>A | r.spl | skeletal, cardiac, GI, GU, other CNS | None | 10-14.99 | MDS; AML | Deceased | 10-14.99 | AML | |
| 40 | 5-9.99 | Asian | No | M | Substitution | Nonsense (stop-gain) | 1 | 36 | c.3601C>T | p.Q1201* | Substitution | Nonsense (stop-gain) | 1 | 36 | c.3601C>T | p.Q1201* | skeletal, GU, eye | 10-14.99 | 10-14.99; 25-29.99 | MDS/AML ; Urothelial carcinoma | Alive | | | 30-34.99 |
| 41 | 15-19.99 | UK | No | M | Substitution | Missense | 0 | 28 | c.2606A>C | p.Q869P | UK | Undetermined | 6 | | | | none | UK | UK | UK | Alive | | | 35-39.99 |
| 42 | 5-9.99 | White | No | F | Substitution | Nonsense (stop-gain) | 1 | 12 | c.1027C>T | p.Q343* | Deletion | Large Deletion | 5 | 1_2 | g.89882258-89883917 | Genomic deletion | skeletal, skin, microcephal[illegible] | 10-14.99 | 25-29.99 | Osteosarcoma | Deceased | 25-29.99 | Cancer | |
| 43 | 5-9.99 | White | No | F | Deletion | Large Deletion | 5 | 4_26 | c.284-?_2504+?del | Genomic deletion | Substitution | Splicing | 4 | i13 | c.1226-2A>G | r.spl | skeletal, skin | 5-9.99 | 25-29.99 | Vulvar SCC, Oral SCC | Alive | | | 25-29.99 |
| 44 | 10-14.99 | White | No | M | Substitution | Splicing | 4 | i21 | c.1827-1G>A | r.spl | Substitution | Splicing | 4 | 21 | c.1827-1G>A | r.spl | skeletal, short stature | 20-24.99 | None | N/A | Alive | | | 20-24.99 |
| 45 | 15-19.99 | White | No | F | Substitution | Splicing | 4 | 36 | c.3624C>T | p.S1208S results in r.spl | Substitution | Nonsense (Stop-gain) | 1 | 12 | c.1027C>T | p.Q343* | skin | UK | UK | UK | Deceased | 25-29.99 | UK | |
| 46 | 1-4.99 | White | No | M | Deletion | Frameshift | 2 | 13 | c.1115_1118delTTGG | p.V372Afs*42 | Substitution | Splicing | 4 | i2 | c.190-2A>T | r.spl | skeletal, skin, GI, GU, other growth, eye | None | None | N/A | Alive | | | 20-24.99 |
| 47 | 10-14.99 | White | No | M | Deletion | Frameshift | 2 | 22 | c.2533_2536delCTCT | p.L845Afs*43 | Substitution | Missense | 0 | 27 | c.1979T>C | p.L660P | skeletal, skin, eye, ear | 15-19.99 | 30-34.99 | Oral cancer | Alive | | | 30-34.99 |
| 48 | 1-4.99 | White | No | M | Deletion | Large Deletion | 5 | 1_43 | g.89722599_89897859del175261 | Genomic deletion | Duplication | Frameshift | 2 | 29 | c.2812_2830dup | p.D944Gfs*5 | skeletal, cardiac, microcephaly, other growth | UK | UK | UK | Alive | | | 15-19.99 |
| 49 | 1-4.99 | White | No | F | Deletion | Frameshift | 2 | 13 | c.1115_1118delTTGG | p.V372Afs*42 | Deletion | Large Deletion | 5 | 16_17 | c.1471-?_1626+?del | Genomic deletion | skeletal, cardiac, GI, GU, eye, ear | 5.2 | None | N/A | Alive | | | 15-19.99 |
| 50 | <1 | White | No | F | Deletion | Frameshift | 2 | 13 | c.1115_1118delTTGG | p.V372Afs*42 | Deletion | Large Deletion | 5 | 16_17 | c.1471-?_1626+?del | Genomic deletion | skeletal, skin, GU, other growth | 1-4.99 | None | N/A | Alive | | | 15-19.99 |
| 51 | 1-4.99 | Asian | No | F | Deletion | Frameshift | 2 | 11 | c.987_990delTCAC | p.H330Afs*4 | Substitution | Splicing | 4 | i2 | c.189+2T>A | r.spl | skeletal, short stature | 10-14.99 | 10-14.99 | MDS | Alive | | | 30-34.99 |
| 52 | 10-14.99 | Black/ African American | No | M | Deletion | Large Deletion | 5 | 13_14 | g.89867339_89876628del9290 | Genomic deletion | Deletion | Large Deletion | 5 | 11_14 | g.89855974_89863563del7590 | Genomic deletion | skin, GU | 10-14.99 | UK | UK | UK | | | 15-19.99 |
| 53 | <1 | White | No | M | Deletion | Large Deletion | 5 | 30_43 | c.2853-?_4368+?del | Genomic deletion | Deletion | Frameshift | 2 | 28 | c.2730_2731delCT | p.W911Dfs*31 | skeletal, cardiac, GI, GU, microcephaly | 6.03 | UK | UK | Alive | | | 15-19.99 |
| 54 | <1 | Other | No | F | Deletion | Large Deletion | 5 | 22_28 | c.1901-?_2778+?del | Genomic deletion | Deletion | Large Deletion | 5 | 22_28 | c.1901-?_2778+?del | Genomic deletion | skeletal, skin, GU, eye, ear | 5-9.99 | 10-14.99 | Liver cancer | Deceased | 10-14.99 | Cancer (liver cancer), liver failure | |
| 55 | 5-9.99 | White | No | M | Deletion | In-frame | 3 | 38 | c.3788_3790delTCT | p.F1263del | Deletion | Large Deletion | 5 | 4_5 | c.284-?_522+?del | Genomic deletion | skeletal, GU, eye | 5-9.99 | 5-9.99 | AML | Deceased | 5-9.99 | Post BMT complications | |
| 56 | 10-14.99 | White | No | F | Substitution | Missense | 0 | 29 | c.2852G>A | p.R951Q | Substitution | Splicing | 4 | i20 | c.1827-1G>A | r.spl | skeletal, skin, GI, microcephaly, short stature, eye, ear | None | None | N/A | Alive | | | 25-29.99 |
| 57 | 10-14.99 | White | No | F | Substitution | Missense | 0 | 29 | c.2852G>A | p.R951Q | Substitution | Splicing | 4 | i20 | c.1827-1G>A | r.spl | skeletal, skin, GI, eye, ear | None | None | N/A | Alive | | | 30-34.99 |

| | | | | | | | | | | | | | | | | | | | | | | | | |
|---|---|---|---|---|---|---|---|---|---|---|---|---|---|---|---|---|---|---|---|---|---|---|---|---|
| 58 | 5-9.99 | White | No | F | Deletion | Frameshift | 2 | 13 | c.1115_1118delTTGG | p.V372Afs*42 | Deletion | Large Deletion | 5 | 9_20 | g.89844987_89869213del24227 | Genomic deletion | skeletal, skin, short stature | 10-14.99 | 10-14.99 | MDS | Alive | | | 20-24.99 |
| 59 | 5-9.99 | White | No | F | Deletion | In-frame | 3 | 38 | c.3788_3790delTCT | p.F1263del | UK | Undetermined | 6 | | | | skeletal, skin, microcephaly, short stature, eye, ear | 5-9.99 | None | N/A | Alive | | | 20-24.99 |
| 60 | 5-9.99 | White | No | F | Substitution | Nonsense (Stop-gain) | 1 | 28 | c.2678G>A | p.W893* | Deletion | Large Deletion | 5 | 16_20 | c.1471_1826del | Genomic deletion | skeletal, GU, other growth, ear | 5-9.99 | None | N/A | UK | | | 20-24.99 |
| 61 | 10-14.99 | White | No | M | Deletion | Large Deletion | 5 | 6_8 | g.89867339_89876628del9290 | Genomic deletion | Deletion | Large Deletion | 5 | 6_8 | g.89867339_89876628del9290 | Genomic deletion | ear | 10-14.99 | UK | UK | Alive | | | 25-29.99 |
| 62 | 10-14.99 | White | No | M | Substitution | Nonsense (Stop-gain) | 1 | 23 | c.2021C>A | p.S674* | Substitution | Missense | 0 | 5 | c.522G>C | p.Q174H | skeletal, skin, other growth, ear | 15-19.99 | UK | MDS; SCC | Alive | | | 30-34.99 |
| 63 | 15-19.99 | White | No | F | Deletion | Large Deletion | 5 | 18_23 | c.1627-?_2778+?del | Genomic deletion | Substitution | Splicing | 4 | i4 | c.426+1G>A | r.spl | none | 15-19.99 | 15-19.99 | Oral SCC | Deceased | 20-24.99 | Cancer | |
| 64 | 10-14.99 | White | No | F | Substitution | Nonsense (Stop-gain) | 1 | 2 | c.154C>T | p.R52* | Duplication | Frameshift | 2 | 29 | c.2815_2816ins19 | p.D944Gfs*5 | skeletal | None | 25-29.99 | Laryngeal SCC | Deceased | 25-29.99 | Cancer | |
| 65 | 15-19.99 | White | No | M | Deletion | In-frame | 3 | 36 | c.3520_3522delTGG | p.W1174del | Substitution | Missense | 0 | 37 | c.3644C>A | p.A1215D | skeletal, skin | 25-29.99 | None | N/A | Deceased | 25-29.99 | Post BMT complications (multiple hemorrhage sites leading to organ failure) | |
| 66 | 1-4.99 | Other | Yes | M | Deletion | Frameshift | 2 | 3 | c.238delT | p.C80Vfs*15 | Deletion | Frameshift | 2 | 3 | c.238delT | p.C80Vfs*15 | cephaly, shor | 1-4.99 | None | N/A | Alive | | | 20-24.99 |
| 67 | 5-9.99 | White | No | F | Substitution | Nonsense (Start-loss) | 1 | 1 | c.1A>T | p.M1? | Substitution | Missense | 0 | 34 | c.3349A>G | p.R1117G | skeletal, skin, GI, GU, short stature | 5-9.99 | None | N/A | Alive | | | 20-24.99 |
| 68 | 5-9.99 | White | No | M | Deletion | Large Deletion | 5 | 13_25 | c.1084-?_2316+?del | Genomic deletion | Deletion | Frameshift | 2 | 30 | c.2872delG | p.A958Rfs*31 | skeletal, skin, eye | 10-14.99 | 25-29.99 | SCC | Alive | | | 30-34.99 |
| 69 | 5-9.99 | Asian | No | M | Substitution | Missense | 0 | 30 | c.2869T>G | p.W957G | Deletion | In-frame | 3 | 30 | g.89820927_89826474del5548 | p.Q952_S994del | skeletal, skin, GU, microcephaly, eye | 10-14.99 | 10-14.99 | Melanoma | Alive | | | 25-29.99 |
| 70 | 5-9.99 | White | Yes | F | Substitution | Missense | 0 | 42 | c.4198C>T | p.R1400C | Substitution | Nonsense (Stop-gain) | 1 | 14 | c.1340C>G | p.S447* | skeletal, skin, short stature | 10-14.99 | None | N/A | Alive | | | 20-24.99 |
| 71 | 1-4.99 | White | No | M | Deletion | Frameshift | 2 | 13 | c.1115_1118delTTGG | p.V372Afs*42 | Substitution | Nonsense (Start-loss) | 1 | 1 | c.2T>C | p.M1? | cardiac, GU, short stature | 5-9.99 | None | N/A | Deceased | 5-9.99 | Post BMT complications (septic episode resulting in multi-organ failure) | |
| 72 | 5-9.99 | white | No | F | Substitution | Missense | 0 | 13 | c.1124T>G | p.L1125W | Substitution | Missense | 0 | 40 | c.4010G>A | p.S1337N | skeletal, skin | 10-14.99 | None | N/A | Deceased | 10-14.99 | Post BMT complications | |
| 73 | 5-9.99 | White | No | M | Substitution | Nonsense (Start-loss) | 1 | 1 | c.2T>C | p.M1? | Substitution | Splicing | 4 | 21 | c.1827-1G>A | r.spl | skin, GU | 15-19.99 | 15-19.99 | MDS | Alive | | | 20-24.99 |
| 74 | 5-9.99 | Other | Yes | M | Deletion | Frameshift | 2 | 4 | c.416_417delTG | p.V129Gfs*41 | Deletion | Undetermined | 6 | 31 | c.2982-?_3066+?del | Genomic deletion | short stature | 5-9.99 | UK | UK | UK | | | 10-14.99 |
| 75 | 5-9.99 | White | No | M | Substitution | Splicing | 4 | i39 | c.3934+1G>A | r.spl | Substitution | Splicing | 4 | i42 | c.4261-2A>G | r.spl | skeletal | None | UK | UK | Alive | | | 20-24.99 |
| 76 | 10-14.99 | Other | No | M | Deletion | In-frame | 3 | 38 | c.3788_3790delTCT | p.F1263del | Deletion | In-frame | 3 | 38 | c.3788_3790delTCT | p.F1263del | skin, other growth | 15-19.99 | 15-19.99 | MDS | Alive | | | 25-29.99 |
| 77 | 10-14.99 | Black/ African American | UK | F | Substitution | Splicing | 4 | i7 | c.710-2A>G | r.spl | Substitution | Splicing | 4 | 28 | c.2778+1G>A | r.spl | skeletal, skin, short stature, other growth | 10-14.99 | None | N/A | Deceased | 25-29.99 | Other (aspiration pneumonia) | |

| | | | | | | | | | | | | | | | | | | | | | | | | |
|---|---|---|---|---|---|---|---|---|---|---|---|---|---|---|---|---|---|---|---|---|---|---|---|---|
| 78 | 5-9.99 | White | No | M | Substitution | Nonsense (Stop-gain) | 1 | 37 | c.3715G>T | p.E1239* | Substitution | Missense | 0 | 28 | c.2639G>A | p.R880Q | skeletal, skin, cardiac, GU, ear | 5-9.99 | 10-14.99 | Pituitary adenoma | UK | | | 10-14.99 |
| 79 | 1-4.99 | White | No | F | Deletion | Large Deletion | 5 | 1_11 | c.-42-?_1006+?del | Genomic deletion | Deletion | Frameshift | 2 | 13 | c.1115_1118delTTGG | p.V372Afs*42 | skeletal, skin, ear | 1-4.99 | None | N/A | Alive | | | 15-19.99 |
| 80 | 1-4.99 | Black/ African American | No | F | Deletion | Large Deletion | 5 | | c.523-?_596+?del | | Substitution | Missense | 0 | | c.542C>T | | skin | 5-9.99 | None | N/A | UK | | | 5-9.99 |
| 81 | 5-9.99 | Other | Yes | F | Deletion | Frameshift | 2 | 11 | c.987_990delTCAC | p.H330Afs*4 | Deletion | Frameshift | 2 | 11 | c.987_990delTCAC | p.H330Afs*4 | skeletal, skin, microcephaly | 10-14.99 | UK | UK | UK | | | 15-19.99 |
| 82 | 1-4.99 | White | No | M | Deletion | In-frame | 3 | 38 | c.3788_3790delTCT | p.F1263del | Substitution | Splicing | 4 | 21 | c.1827-1G>A | r.spl | skeletal, skin, short stature | 5-9.99 | None | N/A | Alive | | | 15-19.99 |
| 83 | 5-9.99 | White | No | F | Substitution | Missense | 0 | 34 | c.3391A>G | p.T1131A | Substitution | Splicing | 4 | i34 | c.3408+1G>A | r.spl | skeletal, skin | 15-19.99 | None | N/A | Alive | | | 20-24.99 |
| 84 | <1 | White | No | M | Substitution | Missense | 0 | 34 | c.3391A>G | p.T1131A | Deletion | Large Deletion | 5 | 1_5 | c.-42-?_522+?del | Genomic deletion | skeletal, cardiac, GU, other CNS | None | None | N/A | Alive | | | 10-14.99 |
| 85 | 1-4.99 | White | No | M | Substitution | Missense | 0 | 28 | c.2738A>C | p.H319P | Deletion | Large Deletion | 5 | 1_29 | c.-42-?_2852+?del | Genomic deletion | skeletal, GU, short stature, eye, ear | 5.34 | UK | UK | Alive | | | 15-19.99 |
| 86 | 1-4.99 | White | No | F | Duplication | Frameshift | 2 | 38 | c.3813dupA | p.H1272Nfs*3 | Deletion | Large Deletion | 5 | 16_17 | chr16(hg19):g.89848927_89850026del | Genomic deletion | skeletal, skin | 5-9.99 | None | N/A | Alive | | | 15-19.99 |
| 87 | 5-9.99 | White | No | F | Deletion | Frameshift | 2 | 13 | c.1115_1118delTTGG | p.V372Afs*42 | Substitution | Missense | 0 | 34 | c.3349A>G | p.R1117G | skeletal, skin, eye | 5-9.99 | None | N/A | Alive | | | 20-24.99 |
| 88 | 1-4.99 | Asian | No | M | Substitution | Splicing | 4 | i40 | c.4010+1G>A | r.spl | Deletion | Large Deletion | 5 | 1_37 | g.89808983_89886611del77629 | Genomic deletion | skeletal, skin, cardiac, GU, microcephaly, short stature, eye, ear | 5-9.99 | 5-9.99 | Pituitary adenoma | Alive | | | 15-19.99 |
| 89 | 1-4.99 | White | No | M | Substitution | Splicing | 4 | i20 | c.1827-1G>A | r.spl | Substitution | Splicing | 4 | i20 | c.1827-1G>A | r.spl | GI, GU, short stature | 1-4.99 | UK | UK | UK | | | 15-19.99 |
| 90 | 5-9.99 | UK | yes | F | Substitution | Missense | 0 | 29 | c.2851C>T | p.R951W | Substitution | Missense | 0 | 29 | c.2851C>T | p.R951W | skin, short stature | None | None | N/A | Alive | | | 15-19.99 |
| 91 | 1-4.99 | White | No | F | Deletion | Frameshift | 2 | 11 | c.916_917delAC | p.T306Afs*32 | Deletion | Large Deletion | 5 | 16_17 | c.1471-?_1626+?del | Genomic deletion | skeletal, skin, cardiac, other growth, eye | 1-4.99 | None | N/A | Alive | | | 15-19.99 |
| 92 | 5-9.99 | White | No | F | Substitution | Splicing | 4 | i41 | c.4168-2A>G | r.spl | Deletion | Large Deletion | 5 | 1_6 | g.89874162_89905652del31491 | Genomic deletion | skin, microcephaly, short stature | 5-9.99 | UK | UK | UK | | | 5-9.99 |
| 93 | 1-4.99 | White | No | F | Substitution | Missense | 0 | 34 | c.3391>G | p.T1131A | Deletion | Large Deletion | 5 | 12_31 | g.89817658_89861785del44128 | Genomic deletion | none | None | None | N/A | Alive | | | 15-19.99 |
| 94 | 5-9.99 | White | No | M | Substitution | Nonsense (Stop-gain) | 1 | | c.1771C>T | p.R591* | Duplication | Frameshift | 2 | 43 | c.4284_4287dupCGAC | p.P1430Rfs*17 | skeletal, skin | 5-9.99 | None | N/A | Alive | | | 15-19.99 |
| 95 | 1-4.99 | Other | No | M | Substitution | Splicing | 4 | i7 | c.710-10G>A | r.spl | Substitution | Splicing | 4 | i7 | c.710-10G>A | r.spl | skeletal, skin, eye | 5-9.99 | None | N/A | Alive | | | 15-19.99 |
| 96 | 5-9.99 | White | No | M | Substitution | Missense | 0 | 34 | c.3391A>G | p.T1131A | Substitution | Splicing | 4 | i28 | c.2778+1G>T | r.spl | skin, GU, short stature | 20-24.99 | None | N/A | Alive | | | 20-24.99 |
| 97 | 5-9.99 | White | No | M | Duplication | Frameshift | 2 | 29 | c.2812_2830dup19 | p.D944Gfs*5 | Deletion | In-frame | 3 | 38 | c.3788_3790delTCT | p.F1263del | other growth, eye | 5-9.99 | None | N/A | Alive | | | 15-19.99 |
| 98 | 5-9.99 | White | No | F | Duplication | Frameshift | 2 | 29 | c.2812_2830dup | p.D944Gfs*5 | Deletion | In-frame | 3 | 38 | c.3788_3790delTCT | p.F1263del | skin | None | None | N/A | Alive | | | 15-19.99 |
| 99 | 5-9.99 | Asian | No | M | Deletion | Undetermined | 6 | 11 | c.894-?_1006+?del | Genomic deletion | Substitution | Nonsense (Stop-gain) | 1 | 32 | c.3188G>A | p.Q1063* | skeletal, skin, short stature | 5-9.99 | None | N/A | Alive | | | 15-19.99 |

| | | | | | | | | | | | | | | | | | | | | | | | | |
|---|---|---|---|---|---|---|---|---|---|---|---|---|---|---|---|---|---|---|---|---|---|---|---|---|
| 100 | 5-9.99 | White | No | F | Deletion | In-frame | 3 | exon 36 | c.3520_3522delTGG | p.W1174del | Substitution | Missense | 0 | 34 | c.3391A>G | p.T1131A | cardiac, GI, microcephaly, short stature, other growth, ear | None | None | N/A | Alive | | | 15-19.99 |
| 101 | 5-9.99 | White | No | M | Substitution | Missense | 0 | 16 | c.1475A>T | p.H492L | Deletion | Large Deletion | 5 | 7_8 | c.597-?_792+?del | Genomic deletion | short stature, eye, ear | 5-9.99 | UK | UK | UK | | | 10-14.99 |
| 102 | 5-9.99 | UK | No | F | Substitution | Splicing | 4 | i9 | c.827-1G>T | r.spl | Deletion | In-frame | 3 | 38 | c.3788_3790delTCT | p.F1263del | skeletal, skin, cardiac, GI, GU | 5-9.99 | None | N/A | Alive | | | 15-19.99 |
| 103 | 5-9.99 | White | No | M | Substitution | Splicing | 4 | i33 | c.3348+1G>A | r.spl | Deletion | In-frame | 3 | 36 | c.3520_3522delTGG | p.W1174del | skin, ear | 10-14.99 | None | N/A | Alive | | | 15-19.99 |
| 104 | 5-9.99 | White | No | F | Deletion | Large Deletion | 5 | 11_14 | c.894-?_1359+?del | Genomic deletion | Substitution | Splicing | 4 | i39 | c.3935-1G>T | r.spl | cardiac, GI, microcephaly, short stature, other growth | 5-9.99 | None | N/A | Alive | | | 15-19.99 |
| 105 | 15-19.99 | White | No | M | Deletion | Frameshift | 2 | 13 | c.1115_1118delTTGG | p.V372Afs*42 | Substitution | Nonsense (Stop-gain) | 1 | 15 | c.1378C>T | p.R460* | skin, GI, short stature | 20-24.99 | UK | MDS | Alive | | | 25-29.99 |
| 106 | 5-9.99 | White | No | M | Deletion | Nonsense (Stop-gain) | 1 | 3 | c.240_241del | p.C80* | Substitution | Missense | 0 | 30 | c.2863C>G | p.H955D | skeletal, ear | 5-9.99 | None | N/A | Alive | | | 15-19.99 |
| 107 | 5-9.99 | Black/ African American | No | F | Substitution | Missense | 0 | 32 | c.3163C>T | p.R1055W | Substitution | Nonsense (Start-loss) | 1 | 1 | c.1A>T | p.M1? | skeletal, cardiac, short stature | 5.92 | None | N/A | Alive | | | 15-19.99 |
| 108 | 1-4.99 | White | No | M | Substitution | Nonsense (Stop-gain) | 1 | 4 | c.295C>T | p.Q99* | Substitution | Missense | 0 | 28 | c.2606A>C | p.Q869P | GI, eye, ear | 10-14.99 | None | N/A | Alive | | | 15-19.99 |
| 109 | 5-9.99 | Other | Yes | F | Deletion | Frameshift | 2 | 3 | c.238delT | p.C80Vfs*15 | Deletion | Frameshift | 2 | 3 | c.238delT | p.C80Vfs*15 | skin, microcephaly, short stature, other growth, eye | None | None | N/A | Alive | | | 15-19.99 |
| 110 | 1-4.99 | UK | Yes | F | Deletion | In-frame | 3 | 38 | c.3788_3790delTCT | p.F1263del | Deletion | Frameshift | 2 | 11 | c.991delA | p.S331Afs*4 | skeletal, cardiac, GI, GU, microcephaly, short stature, other growth, ear | 5-9.99 | None | N/A | UK | | | 5-9.99 |
| 111 | 10-14.99 | White | Yes | F | Deletion | Frameshift | 2 | 30 | c.2910delA | p.G972Afs*17 | Deletion | Frameshift | 2 | 30 | c.2910delA | p.G972Afs*17 | none | 10-14.99 | 10-14.99 | MDS | Deceased | 10-14.99 | Post BMT complications | |
| 112 | 5-9.99 | White | No | M | Substitution | Splicing | 4 | i10 | c.893+1G>T | r.spl | Deletion | Frameshift | 2 | 13 | c.1115_1118delTTGG | p.V372Afs*42 | skeletal, skin | 10-14.99 | None | N/A | Alive | | | 20-24.99 |
| 113 | 5-9.99 | UK | No | M | Substitution | Nonsense (Stop-gain) | 1 | 1 | c.65G>A | p.W22X | Deletion | Frameshift | 2 | 10 | c.890_893del GCTG | p.W298SfsX12 | skin, GU | 5-9.99 | None | N/A | Alive | | | 10-14.99 |
| 114 | 10-14.99 | White | No | F | Deletion | Frameshift | 2 | 37 | c.3696delT | p.F1232Lfs*15 | Deletion | Frameshift | 2 | 37 | c.3696delT | p.F1232Lfs*15 | skeletal, skin, other growth, ear | 10-14.99 | None | N/A | Alive | | | 15-19.99 |
| 115 | 1-4.99 | White | No | F | Substitution | Missense | 0 | 43 | c.4198C>T | p.R1400C | Deletion | Large Deletion | 5 | 1_43 | c.-42_4368+?del | Genomic deletion | skeletal | 5-9.99 | None | N/A | Alive | | | 10-14.99 |
| 116 | 10-14.99 | White | No | M | Substitution | Nonsense (Stop-gain) | 1 | 1 | c.65G>A | p.W22* | Substitution | Splicing | 4 | i39 | c.3935-1G>T | p.D1312Gfs*35 | skeletal, microcephaly, short stature, eye, ear | 20-24.99 | None | N/A | Alive | | | 20-24.99 |
| 117 | 10-14.99 | Other | No | M | Substitution | Splicing | 4 | i42 | c.4261-2A>G | r.spl | Deletion | Large Deletion | 5 | 11_14 | g.89863563_89855974del | Genomic deletion | skeletal, skin | 10-14.99 | None | N/A | Alive | | | 15-19.99 |
| 118 | 1-4.99 | UK | No | M | Deletion | Frameshift | 2 | 13 | c.1115_1118delTTGG | p.V372Afs*42 | Substitution | Nonsense (Stop-gain) | 1 | 19 | c.1771C>T | p.R591* | microcephaly, other growth | 3.4 | None | N/A | Alive | | | 5-9.99 |
| 119 | 5-9.99 | White | No | F | Substitution | Nonsense (Stop-gain) | 1 | 10 | c.862G>T | p.E288* | Deletion | Large Deletion | 5 | not mapped | 16q24.3 | Genomic deletion | skeletal, skin, short stature, other growth, ear | 5-9.99 | UK | UK | Alive | | | 10-14.99 |

| | | | | | | | | | | | | | | | | | | | | | | | | |
|---|---|---|---|---|---|---|---|---|---|---|---|---|---|---|---|---|---|---|---|---|---|---|---|---|
| 120 | 5-9.99 | White | No | M | Substitution | Missense | 0 | 36 | c.3620C>A | p.A1207D | Substitution | Missense | 0 | 36 | c.3620C>A | p.A1207D | skeletal, skin, ear | 5-9.99 | None | N/A | Alive | | | 10-14.99 |
| 121 | 5-9.99 | White | No | M | Substitution | Nonsense (Stop-gain) | 1 | 10 | c.862G>T | p.E288* | Duplication | Frameshift | 2 | 39 | c.3813dupA | p.H1272Tfs*6 | skin, GU, microcephaly, other growth, ear | 5-9.99 | 10-14.99 | Oral SCC | Alive | | | 10-14.99 |
| 122 | 1-4.99 | White | No | M | Substitution | Missense | 0 | 8 | c.710A>G | p.K237R | Substitution | Missense | 0 | 32 | c.3070A>G | p.M1024V | GU, microcephaly, other growth | None | None | N/A | Alive | | | 5-9.99 |
| 123 | 5-9.99 | UK | UK | M | Deletion | Large Deletion | 5 | 1_6 | c.-42-?_596+?del | Genomic deletion | Deletion | Splicing | 4 | i14 | c.1359+1G>C | r.spl | short stature, other growth | 5-9.99 | 5-9.99 ; 10-14.99 | MDS ; Laryngeal SCC | Deceased | 10-14.99 | Cancer | |
| 124 | 5-9.99 | White | No | F | Substitution | Missense | 0 | 29 | c.2852G>A | p.R951Q | Substitution | Splicing | 4 | i33 | c.3349-1G>A | r.spl | skeletal | 10-14.99 | 10-14.99 | Early PTLD | Alive | | | 10-14.99 |
| 125 | 15-19.99 | UK | UK | F | Substitution | Missense | 0 | 28 | c.2639G>A | p.R880Q | Deletion | In-frame | 3 | 38 | c.3788_3790delTCT | p.F1263del | short stature | 25-29.99 | None | N/A | Alive | | | 25-29.99 |
| 126 | 5-9.99 | White | yes | F | Deletion | Large Deletion | 5 | 1_43 | g.89737571_89884589del | Genomic deletion | Substitution | Missense | 0 | 29 | c.2785T>C | p.Y929H | skin, short stature, other growth, ear | 5-9.99 | None | N/A | Alive | | | 10-14.99 |
| 127 | <1 | White | No | M | Deletion | Frameshift | 2 | 13 | c.1115_1118delTTGG | p.V372Afs*42 | Deletion | Large Deletion | 6 | 16_17 | c.1471-?_1626+?del | Genomic deletion | skeletal, GU, short stature | None | None | N/A | Alive | | | 1-4.99 |
| 128 | 10-14.99 | White | No | F | Duplication | Frameshift | 2 | 29 | c.2839dupT | p.S947Ffs*4 | Substitution | Splicing | 4 | i20 | c.1827-1G>A | r.spl | skeletal, skin | None | None | N/A | Alive | | | 15-19.99 |
| 129 | 5-9.99 | White | No | F | Substitution | Splicing | 4 | i16 | c.1566+5G>C | r.spl | Deletion | Large Deletion | 5 | 1_29 | c.-42-?_2852+?del | Genomic deletion | skin, GI, GU, short stature, other growth, ear | None | None | N/A | Alive | | | 10-14.99 |
| 130 | 1-4.99 | White | No | F | Substitution | Missense | 0 | 32 | c.3163C>T | p.R1055W | Substitution | Missense | 0 | 28 | c.2606A>C | p.Q869P | skeletal, skin, other CNS, short stature | None | None | N/A | Alive | | | 5-9.99 |
| 131 | 5-9.99 | White | No | M | Deletion | Frameshift | 2 | 12 | c.1073_1074delTG | p.Y359Pfs*49 | Substitution | Missense | 0 | 40 | c.4004T>C | p.F1335S | skeletal, skin | 5-9.99 | None | N/A | Alive | | | 10-14.99 |
| 132 | 1-4.99 | White | No | F | Deletion | Large Deletion | 5 | 6_20 | c.523-?_1826+?del | Genomic deletion | Substitution | Splicing | 4 | i33 | c.3349-1G>A | r.spl | skeletal, skin, short stature | 1-4.99 | None | N/A | Alive | | | 5-9.99 |
| 133 | 5-9.99 | White | No | F | Duplication | Frameshift | 2 | 29 | c.2812_2830dup19 | p.D944Gfs*5 | Deletion | In-frame | 3 | 38 | c.3788_3790delTCT | p.F1263del | skeletal, skin, short stature, other growth | 5-9.99 | None | N/A | Alive | | | 10-14.99 |
| 134 | 5-9.99 | White | No | M | Deletion | Frameshift | 2 | 12 | c.1074_1075delGT | p.Y359Pfs*49 | Deletion | Frameshift | 2 | 28 | c.2730_2731delCT | p.W911Dfs*31 | skeletal, skin, short stature, other growth | 5-9.99 | None | N/A | Alive | | | 10-14.99 |
| 135 | 5-9.99 | White | No | F | Deletion | Large Deletion | 5 | 1_6 | g.89874512_89884589del | Genomic deletion | Substitution | Nonsense (Stop-gain) | 1 | 10 | c.862G>T | p.E288* | skin, short stature | 5-9.99 | None | N/A | Alive | | | 10-14.99 |
| 136 | 5-9.99 | White | No | M | Substitution | Splicing | 4 | i6 | c.597-1G>C | r.spl | Substitution | Missense | 0 | 27 | c.2527T>G | p.Y843D | skin, cardiac, GI, GU, short stature | 10-14.99 | None | N/A | Deceased | 10-14.99 | Post BMT complications (Renal/ Liver dysfunction, respiratory failure) | |
| 137 | <1 | White | No | M | Deletion | Frameshift | 2 | 41 | c.4015delC | p.L1339Sfs*24 | Deletion | Large Deletion | 5 | 7_10 | c.597-?_827+?del | Genomic deletion | skeletal, GI, short stature | None | None | N/A | Alive | | | 1-4.99 |
| 138 | 1-4.99 | White | No | F | Deletion | Frameshift | 2 | 41 | c.4015delC | p.L1339Sfs*24 | Deletion | Large Deletion | 5 | 7_10 | c.597-?_827+?del | Genomic deletion | skeletal, short stature, ear | None | None | N/A | Alive | | | 5-9.99 |
| 139 | <1 | White | No | M | Deletion | Frameshift | 2 | 41 | c.4124_4125delCA | p.T1375Sfs*49 | UK | Undetermined | 6 | | | | skeletal, skin, GU | 5-9.99 | None | N/A | Alive | | | 5-9.99 |
